# Age-Associated Structural Decline is Linked to Arterial Flow Territories in the Brain: Insights from Lifespan Human Connectome Project in Aging

**DOI:** 10.64898/2026.01.29.26345147

**Authors:** Lindsay C. Hanford, Marziye Eshghi, Jingnan Du, Randy L. Buckner, Ross W. Mair, Tian Ge, Meher R. Juttukonda, David H. Salat

**Affiliations:** Department of Psychology, Center for Brain Science, Harvard University, Cambridge, MA 02138, USA; Athinoula A. Martinos Center for Biomedical Imaging, Massachusetts General Hospital, Charlestown, MA 02129, USA; MGH Institute of Health Professions, Charlestown, MA, USA; Department of Psychiatry, Massachusetts General Hospital, Charlestown, MA 02129, USA; Department of Radiology, Harvard Medical School, Massachusetts General Hospital, Boston, MA, USA; Psychiatric and Neurodevelopmental Genetics Unit, Center for Genomic Medicine, Massachusetts General Hospital, Boston, MA, 02114, USA; Center for Precision Psychiatry, Massachusetts General Hospital, Boston, MA, 02114, USA; Stanley Center for Psychiatric Research, Broad Institute of MIT and Harvard, Cambridge, MA, 02138, USA; Neuroimaging Research for Veterans (NeRVe) Center, VA Boston Healthcare System, Boston MA 02130

**Keywords:** Aging, brain structure, cortical thickness, hippocampus, hemodynamics, MRI, ASL, cerebral blood flow, arterial transit time

## Abstract

The effect of biological aging on brain structure is widespread and apparent. However, little is understood regarding which regions exhibit similarities in vulnerability, and what biological processes drive regional patterns of senescence-associated atrophy. Here, we investigated whether associations between age and brain structure exhibit distinct patterns of regional vulnerability, and if so, whether they are related to patterns of cerebral physiology which also show age-related decline. Using both data-driven and hypothesis-driven approaches, we identified recurring patterns of *accelerated* and *delayed* decline across the lifespan. Notably, the results mapped using unsupervised clustering methods mirrored the organization of major arterial flow territories, suggesting that vascular architecture may serve as a key organizing principle in brain aging. These results provide support for future research on aging and neurodegenerative disorders that aim to link patterns of structural deterioration to physiological processes that may be useful for identifying ‘at risk’ individuals and developing novel therapeutics.

## Introduction

The effect of aging on the brain is robust. Widespread reductions in brain volume and cortical thickness are apparent from postmortem examinations of brain tissue ^1–3^ as well as from in vivo studies using noninvasive magnetic resonance imaging (MRI) techniques [to list a few: ^4–18^]. Yet, regional analyses reveal heterogeneous rates of decline, ranging from approximately 0.2–2% annually ^8–10^. Previous work investigating regional patterns with age have observed the strongest and/or most *accelerated* rates of decline in the medial prefrontal, posterior cingulate, precuneus, and along superior frontal and superior temporal cortices ^9,13,14,19,20^. Meanwhile, medial temporal regions including entorhinal, parahippocampal, as well as lateral prefrontal and occipital cortices ^7,9,10,12,14,20^ are relatively spared. Other regions, including the hippocampus, show a *delayed* decline with age (i.e. a nonlinear relationship where atrophy accelerates with age) ^8,10,21–23^. These age associations have been observed and reported in multiple cross-sectional and longitudinal cohorts, and support regionally specific patterns of *accelerated* and *delayed* decline across the cortex. While it is clear regional heterogeneity exists, the biological principles underlying these spatial patterns remain poorly understood.

One potential organizing factor shaping this heterogeneity could be the brain’s vascular architecture. Arterial flow territories, defined by regions supplied by the anterior, middle, and posterior cerebral arteries, represent an intrinsic topographical framework that could influence both developmental trajectories and vulnerability to age-related change. These territories differ in perfusion characteristics, oxygen supply, and vascular risk exposure, which may in turn influence the rate and pattern of structural or functional decline ^24–29^. Recent high-resolution vascular imaging and animal work has illustrated that aging drives cerebrovascular remodeling, including reductions in vessel length and branching, and increased vascular tortuosity, particularly within deep cortical structures and the hippocampus ^30^. In parallel, human studies have highlighted that systemic vascular aging may manifest as arterial stiffening, endothelial dysfunction, and impaired angiogenic response, and play a central role in neural vulnerability ^31^. These findings suggest that vascular topology may be a foundational scaffold constraining regional rates of decline through its effects on perfusion, oxygen delivery, and metabolic support.

Given that neuronal maintenance is energetically expensive and strictly dependent on perfusion, regional differences in vascular architecture could explain why certain regions age more rapidly than others. Importantly, the brain’s arterial territories are established during development and remain stable through adulthood, implying that early vascular organization may influence later regional susceptibility. Yet, despite this conceptual link, prior studies have not systematically tested whether the spatial pattern of age-related brain decline aligns with the brain’s arterial topography.

The use of arterial spin labeling (ASL) magnetic resonance imaging has gained popularity over the past 20 years for evaluating cerebral microvascular physiology ^32–35^. ASL imaging can provide quantitative measures of cerebral blood flow (CBF) and arterial transit time (ATT), which represent sensitive markers of perfusion and vascular transit dynamics ^35–37^. Previous work has reported both global and regional age-related decline in CBF and increase ATT ^38–40^, consistent with diminished vascular efficiency.

In this study, we investigated whether associations between age and brain structure exhibit distinct patterns of regional vulnerability, and if so, whether they are related to patterns of cerebral physiology which also show age-related decline. We hypothesized that brain regions sharing a common arterial supply would exhibit correlated trajectories of age-related structural decline, reflecting the organizing role of vascular topology. By integrating structural MRI and ASL-derived perfusion metrics acquired within individuals, we aimed to identify whether physiological organization along arterial flow territories contribute to the spatial patterning of age-related brain change. To this end, we first evaluated whether certain brain regions “*age*” together, and second, whether these structural patterns of *accelerated* and *delayed* decline were topologically organized in a manner that matches the vascular territories of the brain.

## Methods

### Overview

To assess which brain regions age together, regional age-associations were estimated from the Lifespan Human Connectome Project in Aging (HCP-A) dataset ^72,73^ and arranged using an unsupervised clustering algorithm. To reduce the influence of noise in our model, prior to clustering, regions observed to have low test-retest reliability in an independent sample were excluded (R^2^ < 0.6; see *Supplemental Materials 2* for full details). To confirm these age patterns were not an effect of cohort, a subset of structural estimates were employed as an independent replication from the UK BioBank ^74^. Age-associations with brain structure and brain physiology were compared in two ways: (1) First, we compared the spatial topology of age associations clustered by likeness, and uninformed by vascular organization, with ones defined by vascular flow territories. (2) Second, age associations from estimates obtained from T1-weighted MRI were compared to those derived from ASL images in the same individuals.

### Human Connectome Project-Aging (HCP-A)

#### Participants

Participants included in this study consisted of 830 healthy adults between 36-89 years of age. Demographic, biological, and behavioral information is summarized by age in Table 1. Of the full HCP Aging dataset, 101 participants were excluded for poor T1-weighted image quality and 219 for unavailable or missing covariable data. Finally, as our reliability were estimated based on sample size of 30 or greater, sample size for individuals above 90 years (n = 25) had insufficient power and for this reason were excluded. Sex, years of education, and group sizes were well balanced across age. A subset of individuals from the HCP-A cohort participated in the collection of both T1-weighted and ASL images (n = 656; see Supplementary Table 1).

**Table 1.** HCP Aging sample demographics and performance scores on cognitive tasks across the adult lifespan (36-89 years).

|  | 36-39 | 40-44 | 45-49 | 50-54 | 55-59 | 60-64 | 65-69 | 70-74 | 75-79 | 80-84 | 85-89 |
| --- | --- | --- | --- | --- | --- | --- | --- | --- | --- | --- | --- |
| n | 73 | 92 | 86 | 81 | 90 | 72 | 69 | 72 | 67 | 72 | 56 |
| % Female | 51 | 38 | 41 | 41 | 42 | 47 | 46 | 50 | 49 | 46 | 45 |
| Education in Years,<br>Mean (SE): | 17.43<br>(0.28) | 17.51<br>(0.24) | 17.58<br>(0.23) | 17.16<br>(0.25) | 16.86<br>(0.25) | 17.86<br>(0.20) | 17.84<br>(0.25) | 17.63<br>(0.26) | 17.76<br>(0.29) | 17.25<br>(0.31) | 17.23<br>(0.32) |
| MOCA Summary Score,<br>Mean (SE): | 26.99<br>(0.26) | 26.82<br>(0.24) | 26.76<br>(0.27) | 26.32<br>(0.29) | 26.14<br>(0.27) | 26.67<br>(0.29) | 26.38<br>(0.29) | 26.69<br>(0.27) | 25.61<br>(0.31) | 25.21<br>(0.32) | 23.79<br>(0.36) |
| TrailA RT,<br>Mean (SE): | 26.81<br>(1.40) | 26.12<br>(1.10) | 25.24<br>(0.09) | 26.41<br>(1.12) | 28.41<br>(1.01) | 27.91<br>(1.30) | 33.35<br>(1.39) | 31.87<br>(1.21) | 38.51<br>(1.46) | 41.60<br>(1.74) | 44.26<br>(2.47) |
| TrailB RT,<br>Mean (SE): | 63.76<br>(3.02) | 63.97<br>(3.93) | 62.10<br>(3.75) | 65.65<br>(3.99) | 69.83<br>(3.99) | 69.08<br>(4.05) | 83.26<br>(5.17) | 78.86<br>(5.41) | 96.46<br>(5.38) | 107.53<br>(9.56) | 122.61<br>(9.12) |

RT = Reaction Time; SE = Standard Error

#### Data Acquisition

Whole-brain T1- and T2-weighted MRI images acquired using 32-channel head coil and 3-Tesla Siemens Prisma scanners across four study sites. Details on the HCP-Aging imaging acquisitions protocols have been described previously ^72,73,75,76^, and all HCP-style pulse sequences available for distribution at http://protocols.humanconnectome.org/lifespan/. The T1-weighted MRI was acquired using a multi-echo magnetization-prepared rapid gradient echo (MPRAGE) sequence ^77^ with the following sequence parameters: TR = 2500ms, TI = 1000ms, TE (4 echoes) = 1.8/3.6/5.4/7.2ms, flip angle = 8 deg, resolution = 0.8 mm isotropic, scan duration = 8:22 minutes (maximum). T2-weighted images were acquired using a turbo-spin-echo (TSE) sequence (Mugler III et al. 2000) with the following parameters: TR = 3200ms, TE = 564ms, turbo factor = 314, resolution = 0.8 mm isotropic, scan time = 7:45 minutes (maximum).

Pseudo-continuous ASL (PCASL) images were acquired without background suppression, and with labeling duration of 1500ms, and five post-labeling delays (PLD) = 200ms (control/label pairs = 6), 700ms (pairs = 6), 1200ms (pairs = 6), 1700ms (pairs = 10), and 2200ms (pairs = 15) ^78^. Signal readout was performed using multi-band 2D gradient echo echo-planar imaging (EPI) sequence with the following parameters: TR = 3580ms; TE = 19ms; partial Fourier = 6/8; SMS acceleration factor = 6; total slices = 60; resolution = 2.5 mm isotropic, scan time = 5.5 min. Two equilibrium magnetization (M0) images were acquired at the end of the scan. A pair of spin echo EPI scans with opposite phase encoding directions were also acquired for use in the correction of field inhomogeneity-related distortions: TR = 8000ms; TE = 40ms; spatial resolution = 2.5 mm isotropic.

#### Image Preprocessing

Structural images were preprocessed using HCP-A’s minimal processing pipeline prior to use ^76^. Full details have been described in detail previously ^73,75,76,79^. Briefly, T1-weighted and T2-weighted images were refaced and corrected for gradient-field, bias-field, and readout-related distortions and aligned in native T1-weighted image space using FreeSurfer’s Imaging Suite (v6.0.0; https://freesurfer.net/). Automated segmentation of subcortical structures ^80^, including: the amygdala (AMYG), caudate (CAUD), hippocampus (HIPP), globus pallidus (PALL), putamen (PUT), and thalamus (THAL), ventral diencephalon (vDC), and the Accumbens or Ventral Striatum (VS) were used to provide estimates of brain structure. Regional cortical estimates were obtained from reconstructed 3D cortical surfaces ^81^, and defined using the Desikan-Killiany atlas ^82^. This parcellation scheme is already widely used, with regional estimates are ready for use in many publicly available datasets (including HCP-A and UKBiobank), and provides a common space for across imaging modalities. Additionally, this parcellation has previously demonstrated high validity with manual tracing, a high degree of reliability across repeated scanning, and high sensitivity for detecting brain atrophy associated with healthy aging and neurodegenerative disorders ^82^. The atlas includes regions (and abbreviation): caudal and rostral Anterior Cingulate Cortex (cACC, rACC), Peri<u>cal</u>carine sulcus (CAL), <u>Cun</u>eus and <u>Precun</u>eus Cortex (CUN, PreCUN), Entorhinal Cortex (EC), Fusiform Gyrus (FG), Frontal Pole (FP), Isthmus Cingulate Cortex (ICC), pars opercularis (IFGoperc), pars orbitalis (IFGorb) and pars triangularis (IFGtri) of the Inferior Frontal Gyrus, Insular Cortex (Ins), Inferior Parietal Cortex (IPC), Interior Temporal Gyrus (ITG), Lingual Gyrus (LG), Lateral Occipital Cortex (LOC), caudal and rostral Middle Frontal Gyrus (cMFG, rMFG), Middle Temporal Gyrus (MTG), lateral and medial Orbital Frontal Cortex (lOFC, mOFC), <u>Parac</u>entral Lobule (ParaC), <u>Pre</u>- and <u>Po</u>st-<u>c</u>entral Gyrus (PreC, PoC), Posterior Cingular Cortex (PCC), Parahippocampal Gyrus (PHG), Superior Frontal Gyrus (SFG), Supramarginal Gyrus (SMG), Superior Parietal Cortex (SPC), bank of Superior Temporal Sulcus (bSTS), Superior Temporal Gyrus (STG), Temporal Pole (TP) and Transverse Temporal Cortex (TT). Regions observed to have low test-retest reliability in an independent sample included the mOFC, lOFC, INS, PreC, bankSTS, rACC, and TP were excluded (R^2^ < 0.6; see *Supplemental Materials 2* for more details). Additionally, while caudate volume appeared reliable, it was excluded as associations with age had low biological interpretability (see *Supplemental Materials 2*). These regions were excluded to reduce the influence of noise in our model prior to clustering.

ASL images were preprocessed using a pipeline described previously ^38,40^. Briefly, ASL data were corrected for susceptibility distortions using the spin echo EPI data and TopUp from FSL ^83,84^. Motion correction was performed using in-house routines. Control and label ASL data were pair-wise subtracted, averaged for each post labeling delay, and normalized to the second M0 image. Correction for partial volume effects was performed using PET Surfer ^85^.

#### Arterial Transit Time

ATT values were calculated on a voxel-wise basis through a two-stage normalized cross-correlation approach described and validated previously ^38^. First, the signal time course in each voxel was cross-correlated with that of a simulation from the flow-modified Bloch equation with 11 shift indices ranging between −5 to +5 ^38^. The maximum correlation between the acquired and simulated time courses and the corresponding time shift were recorded. ATT values were assigned based on a second cross-correlation analysis where the measured and simulated time courses were again cross-correlated, this time, with temporal resolution = 0.1 s for ATTs between 0.2 −3.2 s. Here the maximum correlation defined for each shift index was used as the ATT.

#### Cerebral Blood Flow

CBF values were calculated using a two-compartment model described previously ^38–40,86^. Longitudinal relaxation rates of arterial blood needed for this computation were derived from hematocrit values ^87^ that were estimated on an individual basis using a previously characterized relationship appropriate for age and sex ^88^. A final CBF map was derived using a least-squares fitted model that included data from all PLDs and ATT values estimated at each voxel ^40^.

### Arterial Flow Territories

To explore whether age-related patterns in structure across the brain align with the topography of arterial supply, a map of the arterial flow territories of the cortical surface from the three main arterial branches: anterior (ACA), middle (MCA) and posterior (PCA) cerebral arteries, available for download on NITRC was employed (https://www.nitrc.org/projects/arterialatlas/). This atlas was created by averaging data from 1,298 patients who experienced acute and subacute ischemic strokes ^89^. Automated cortical regional parcellations were assigned to one of three arterial flow territories based the majority overlap (i.e. the # of voxels/vertices of a given region assigned to the territory > 70%). Three regions: the precuneus, superior parietal, and isthmus cingulate cortex, fell nearly equally along two or three arterial flow territories and were excluded from this analysis.

### UK BioBank (Replication Cohort)

#### Participants

Our study made use of estimates generated by an image-processing pipeline developed and run on behalf of the UK Biobank ^90^. Participant details have been described previously ^74^. Briefly, subjects were 35334 healthy adults between 44-82 years of age that participated across the UK. Of the full UK BioBank sample (n = 42184), subjects were excluded due to data missingness (n = 555), Body mass index (BMI) exceeded 40 (n = 327), or the image quality was not reported (T1 invSNR n = 5150) or poor (n = 818). Sex and years of education, and size of groups were well balanced across age (see Table 2).

**Table 2.** UK BioBank sample demographics and performance scores on cognitive tasks across the adult lifespan (44-82 years).

|  | **44-49** | **50-54** | **55-59** | **60-64** | **65-69** | **70-74** | **75-82** |
| --- | --- | --- | --- | --- | --- | --- | --- |
| n | 843 | 4460 | 5849 | 7235 | 8289 | 6384 | 2274 |
| % Female | 45 | 44 | 42 | 44 | 48 | 53 | 58 |
| % Right-Handed | 89 | 89 | 89 | 89 | 89 | 89 | 90 |
| Education in Years,<br>Mean (SE): | 17.04<br>(0.14) | 17.25<br>(0.06) | 17.31<br>(0.05) | 17.20<br>(0.05) | 17.02<br>(0.04) | 16.8<br>(0.04) | 16.6<br>(0.08) |
| TrailA RT,<br>Mean (SE): | 18.42<br>(0.35) | 19.32<br>(0.11) | 20.03<br>(0.09) | 21.06<br>(0.10) | 22.74<br>(0.12) | 25.00<br>(0.15) | 27.36<br>(0.32) |
| TrailB RT,<br>Mean (SE): | 42.98<br>(0.88) | 46.17<br>(0.32) | 48.29<br>(0.31) | 51.50<br>(0.31) | 56.67<br>(0.37) | 62.94<br>(0.50) | 67.70<br>(0.92) |

RT = Reaction Time; SE = Standard Error

#### Imaging Acquisition and Processing

Structural scans were acquired at 3-Tesla on a Siemens Skyra with 32-channel head receive coil. T1-weighted imaging was obtained using a 3D MPRAGE sequence ^91^ with the following parameters: TR = 2000ms, TI = 880ms, flip angle = 8 deg, voxel size = 1.0 mm isotropic, scan time = 4:54 minutes. T2 fluid-attenuated inversion recovery (FLAIR) imaging ^92^ was also acquired with TR = 5000 ms, TI = 1800 ms, resolution = 1 x 1 x 1.05 mm elliptical, scan time = 5:52 minutes. Images were pre-normalized during acquisition. Raw, refaced T1-weighted images (and T2_FLAIR images when possible) were processed using standard FreeSurfer methods described in previous publications ^21,90^.

### Statistical Analyses

All statistical analyses were performed in R https://www.r-project.org/ (version 4.0.2; https://www.r-project.org/), utilizing packages including reshape2, plyr, Hmisc and ggplot2 for visualizations.

#### Adjusting estimates for differences in head size

Estimated Total Intracranial Volume (eTIV), commonly used to adjust for differences in head size between individuals, was found to be correlated with age in the HCP dataset. For this reason, raw measures of eTIV were adjusted with an atlas transform error score (available through FreeSurfer) using a method published previously (see *Appendix 1* or ^23^). All volume-based estimates, as well as ASL-based estimates have been adjusted for differences in head size using this corrected measure of eTIV (see *Supplemental Materials 1*).

#### Normalizing measures across the lifespan

To compare associations with age, brain estimates from participants 40 years and older were compared to those from the youngest age range (i.e. 36-39 years), as the percent difference across age. Individual estimates were pooled into five-year age bins, and the mean of each age bin was used to calculate the normalized percent difference of that group relative to the youngest group. Where applicable, the percent annualized rate (PAR) – calculated as the percent difference per year (i.e. %/year) – is reported. Normalized estimates provide several benefits, including enabling comparisons across different units of measures (e.g. thickness vs. volume), or across dataset where sample-based biases may be present. Finally, by anchoring to a fixed start point (i.e. estimates of the 1^st^ age bin) all regions share the same initial value (100%), allowing comparisons of “shape” or magnitude of slope to be easily interpretable of the age associations between regions. Finally, as ATT is the only measure expected to increase with age, estimates of normalized ATT have been inverted for ease of comparison to all other age-associations that decrease with age.

#### Unsupervised hierarchical clustering methods

To investigate whether estimates shared similar patterns of aging across the cortex, an unsupervised hierarchical clustering algorithm was employed using R package *hclust*. The normalized age association curve for each region was used as the input to the model. Each region was assigned to a group by computing the Euclidian distance of its age association and applying Ward D2 algorithm ^93^ with respect to each other regions age-association normalized curve.

## Results

### Regional structural estimates show distinct patterns of accelerated, steady and delayed decline with age

Using data from the HCP Aging cohort, regional subcortical volumes clustered into four distinct patterns with age across the lifespan (see Figure 1). Figure 1A displays the results of regions grouped by likeness based on their age associations as a heatmap, and a branch visualization of the hierarchical clustering. Figure 1B displays the average age curve of all regions assigned to each group, while 1C displays each region’s age-association curve within each of the assigned groups. The first group consisted of the ventral striatum and was observed to have the steepest or most *accelerated* pattern of decline with age compared to other subcortical regions (VS: −0.74 %/year)(Figure 1Bi). The second group consisted of the brainstem (BS: −0.16%/year) and showed little-to-no decline, or *maintained* structure across the lifespan (Figure 1Bii). The third group showed *delayed* decline (i.e. minimal decline early in adulthood and accelerated decline later in adulthood)(Figure 1Biii). This group included the hippocampus (HIPP) and globus pallidus (PALL). Finally, regions assigned to the fourth cluster demonstrated *steady* decline with age and included the ventral diencephalon (vDC), putamen (PUT), amygdala (AMYG), and thalamus (THAL)(Figure 1Biv).

**Figure 1.**
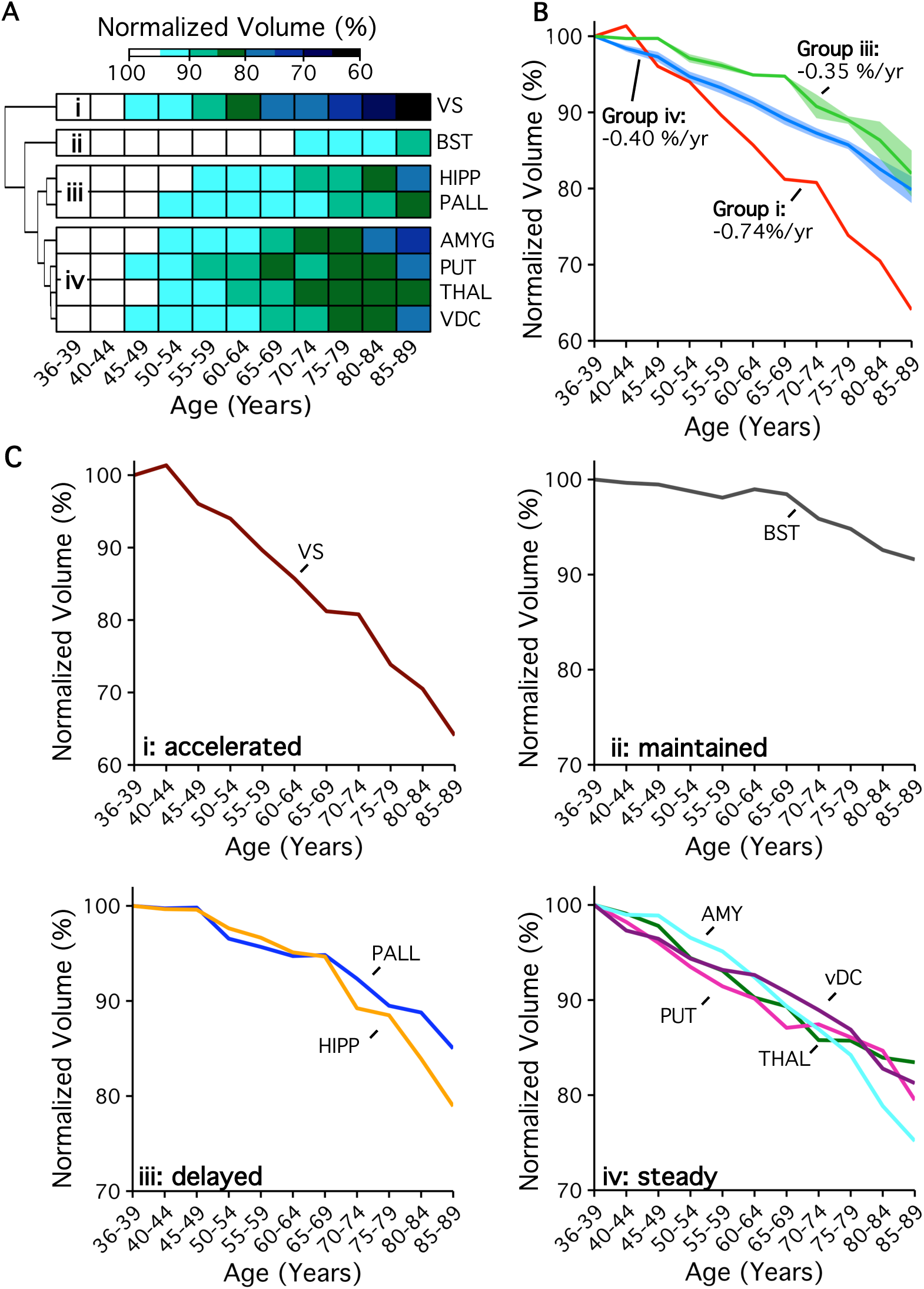
Regional subcortical volumes followed four distinct patterns across the lifespan. (A) Regional age associations demonstrated patterns of (i) accelerated, (ii) maintained, (iii) delayed; and (iv) steady decline across adulthood. Regional age associations are arranged by likeness and visualized using a heatmap, and clustering visualized with branches. For all regions, the normalized difference (%) was estimated to that of the first age group. (B) Accelerated, steady and delayed decline cluster mean age associations are displayed on one plot, and (C) each plot displays individual regional age associations assigned to each group.

Regional cortical thickness followed similar patterns of *delayed*, *steady*, and *accelerated* decline across the lifespan (see Figure 2). The first group demonstrated *delayed* decline and included the frontal pole (FP), cuneus (CUN), fusiform gyrus (FG), inferior temporal gyrus (ITG), entorhinal cortex (EC), lateral occipital cortex (LOC), pars orbitalis of the inferior frontal gyrus (IFGorb), lingual gyrus (LG) and parahippocampal gyrus (PHG) (Figure 2Bi). Most of the cortex was assigned to the second group, which demonstrated a pattern of *steady* linear decline across the lifespan (Figure 2Bii, also see Methods for a full list of regions and their abbreviations). Finally, the third group which included caudal anterior cingulate cortex (cACC), posterior cingulate cortex (PCC), isthmus cingulate cortex (ICC), and transverse temporal (TT) regions followed a pattern of *accelerated* decline with age (Figure 2Biii). The average age curve of all regions assigned to each group can be seen in Figure 2C. The annual rate of decline was almost double for the *accelerated* group compared to the *delayed* decline group on average. It is worth noting that shared age patterns of *accelerated*, *steady* and *delayed* decline were found across subcortical regions as well as for cortical regions even from clustering independently.

**Figure 2.**
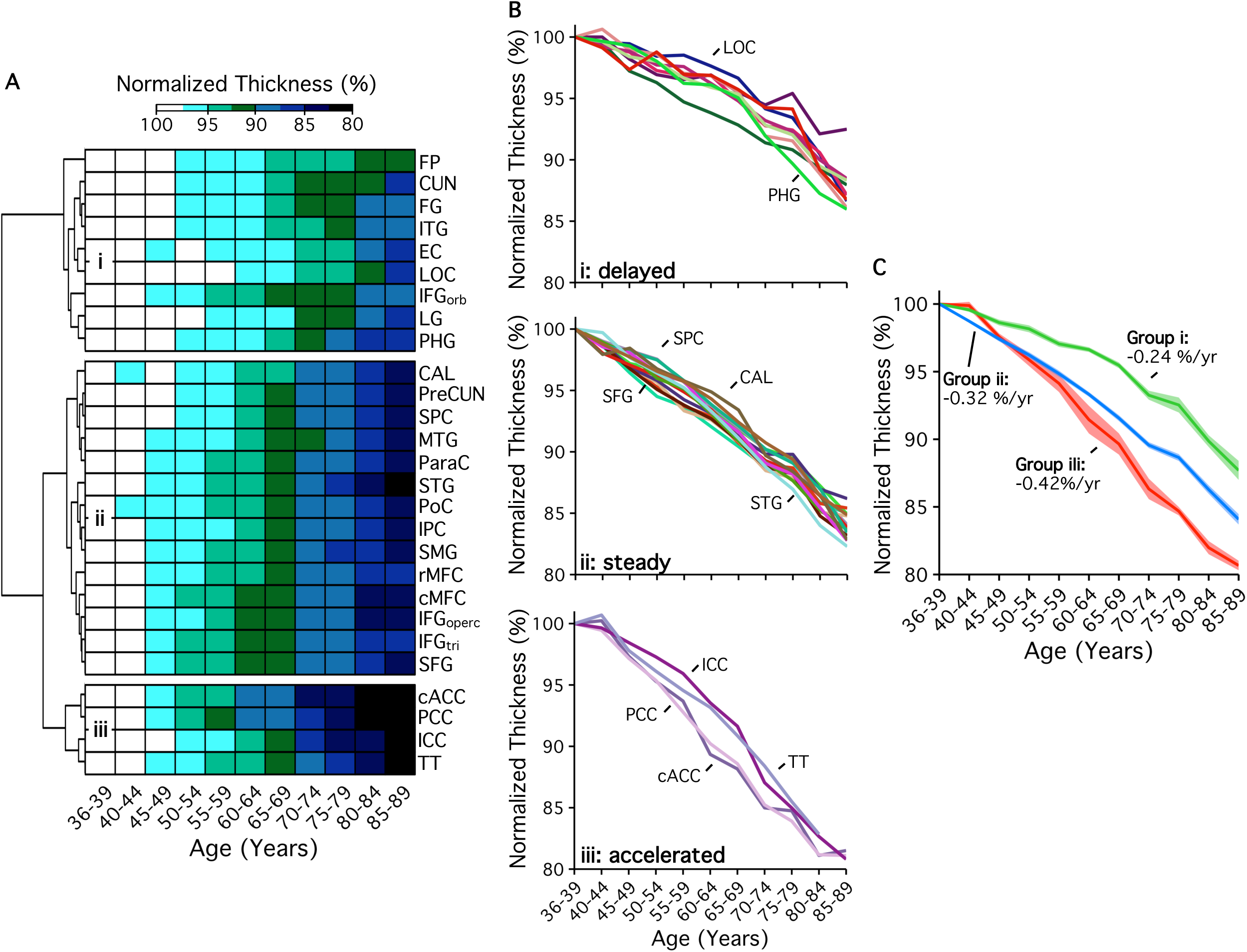
Regional cortical thickness followed three distinct patterns across the lifespan. Regional age associations demonstrated patterns of (i) accelerated, (ii) delayed, and (iii) steady decline across adulthood. As with Figure 1, Regional age associations are arranged by likeness and visualized using a heatmap, and clustering visualized with branches. For all regions, the normalized difference (%) was estimated to that of the first age group. (B) each plot displays individual regional age associations assigned to each group, and (C) Accelerated, steady and delayed decline cluster mean age associations are displayed on one plot.

### Age patterns of accelerated, steady, and delayed decline map to arterial flow territories of the brain

To determine whether age patterns of *accelerated, steady, delayed* decline share common topology with brain physiology, we compared the results from our data-driven unsupervised clustering approach (left side) to assignment based on one of three cerebral arterial flow territories (right side); the anterior cerebral artery (ACA), middle cerebral artery (MCA), or posterior cerebral artery (PCA) (Figure 3). Regions excluded due to low reliability or not assigned to a flow territories have been described in the methods. While most subcortical structures are supplied by two or more arterial branches, the VS is the only subcortical structure explored here that is predominantly supplied by the ACA ^41,42^. The vDC, THAL and HIPP are predominantly supplied by the PCA ^25,43–45^, while the PALL, PUT, and AMYG are supplied by the MCA ^46–48^. Next, for cortical regions, group assignment between data-driven and hypothesis-based approaches was compared using percent agreement between each of the assigned groups. Using a data-driven, unsupervised clustering approach, cortical regions were classified into groups that reflect flow territories with overall 83.3% agreement: with regional classification to the ACA having 87.5% agreement, the MCA with 75.0%, and PCA with 87.5% agreement. These values support measurable overlap between cortical structural integrity and vascular architecture.

**Figure 3.**
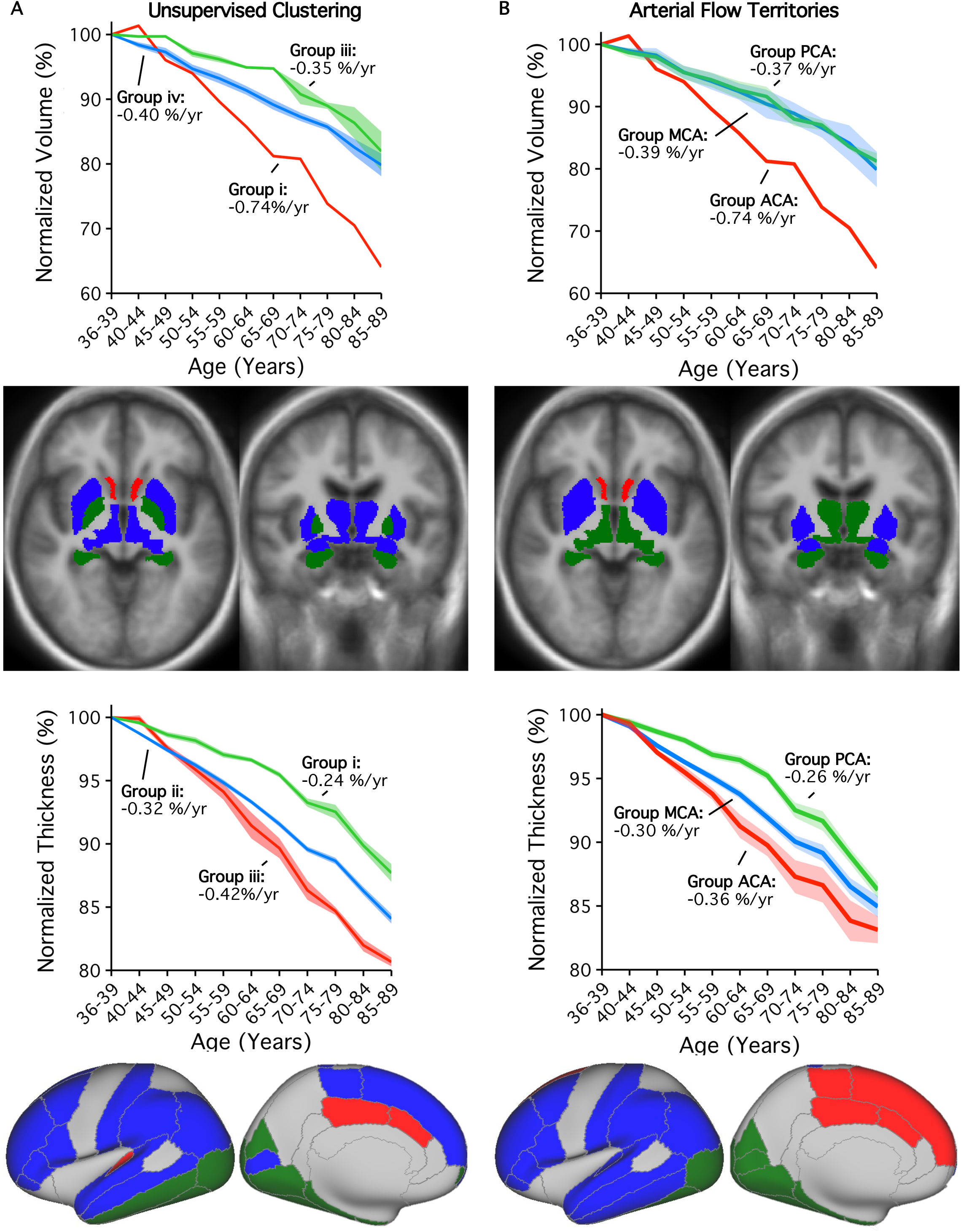
Age patterns mapped to known arterial flow territories of the brain. Side-by-side we compare the spatial topology of results from (A) unsupervised clustering model to (B) those defined by known arterial flow territories: Anterior (ACA), Middle (MCA), and Posterior (PCA) Cerebral Arteries of the brain. Patterns of accelerated and steady decline are seen when clustering subcortical normalized volumes by arterial flow territories. Patterns of accelerated, steady and delayed decline are seen when grouping regional cortical thickness by arterial flow territories.

### Age patterns were replicated in an independent sample

To support that these results are not an effect of cohort, age patterns were further explored using data from 35334 participants from the UK BioBank sample. In Figure 4, the top row displays our previous observations from the HCP Aging cohort of regional subcortical (i.e. HIPP volume) and cortical patterns of *delayed* (i.e. LOC and PHG thickness), and *accelerated* decline (i.e. cACC and PCC thickness). Despite the slightly narrowed window across the adult lifespan (44-82 yrs), nearly all regional age patterns of decline replicated. The UK Biobank cohort also demonstrated *delayed* decline in hippocampal volume, as well as LOC and PHG thickness compared to the rest of the cortex. Patterns of *accelerated* decline replicated for cACC, but not PCC thickness. The age association found with PCC thickness was comparable to that of the average across the cortex.

**Figure 4.**
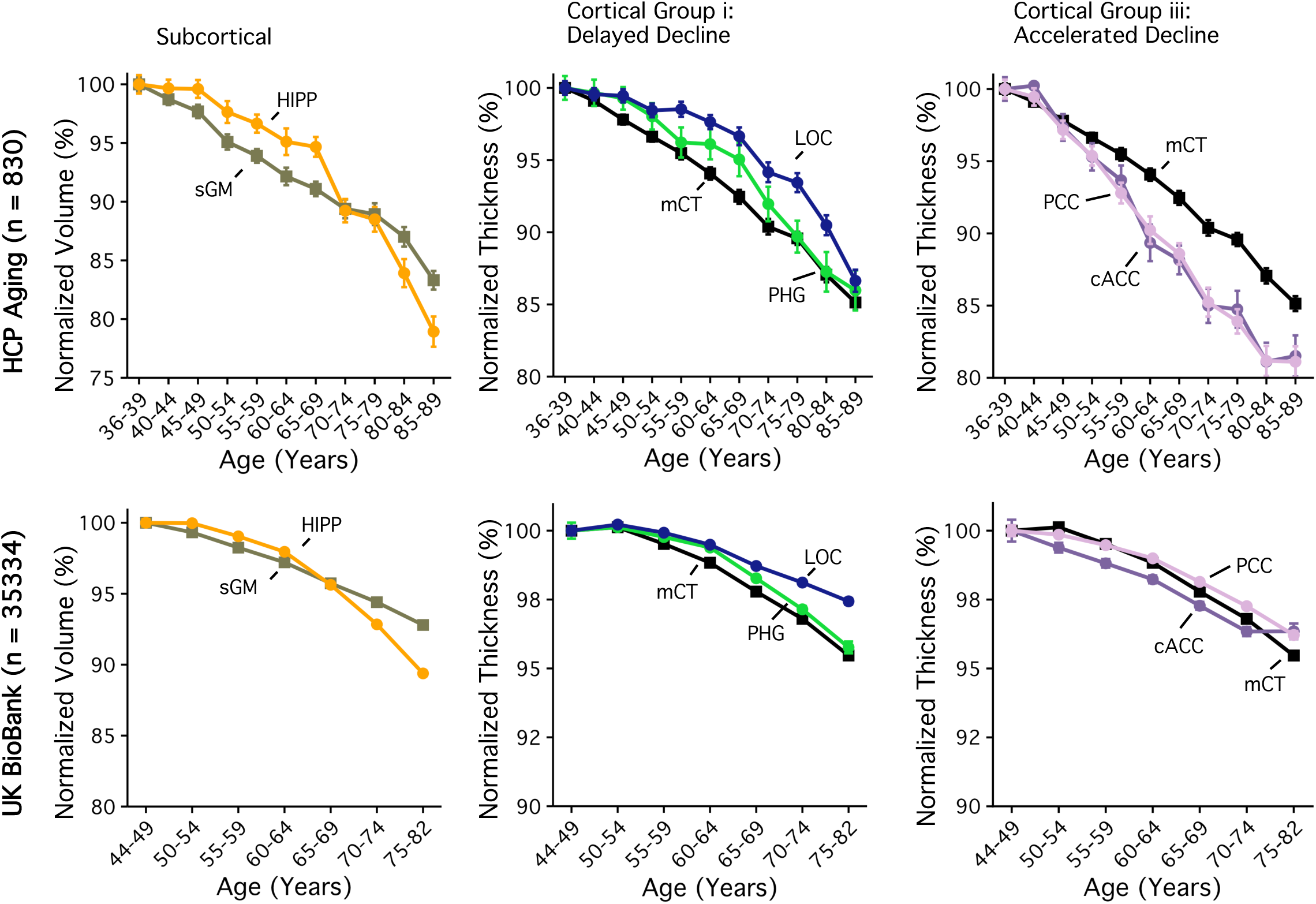
Age patterns delayed and accelerated decline across subcortical and cortical regional estimates were replicated in an independent sample. Regional subcortical (i.e. HIPP volume) and cortical patterns of *delayed* (i.e. LOC and PHG thickness), and *accelerated* decline (i.e. cACC and PCC thickness) were displayed using data from the HCP Aging cohort (top row), and replicated using the UK BioBank cohort (bottom row). Despite a narrowed window across the adult lifespan (44-82 yrs), with the exception of the PCC, all regional age patterns were replicated. Normalized difference (%) was estimated to that of the first age group.

### Estimates from ASL images show distinct age patterns

Building on our previous work of characterizing cerebral blood flow (CBF) and arterial transit time (ATT) in a subset of the HCP-Aging cohort from MGH ^38,39^, Figure 5 displays regional associations with age in the expanded sample (see Supplemental Materials 2 for associations with global cortical gray matter, white matter, and subcortical gray matter). Our previous observations that global tissue types carry differential age associations across T1-weighted and ASL images provided support to explore regional age associations of CBF and ATT that displayed robust age associations using T1-weighted MRI. In Figure 5 we display the same set of key regions representing *delayed* (i.e. HIPP volume, LOC and PHG thickness), and *accelerated* decline (i.e. PCC and cACC) with age. The top row displays the same volume and thickness based normalized age curves seen in previous figures, while the middle and bottom rows show the CBF and ATT in these same regions.

**Figure 5.**
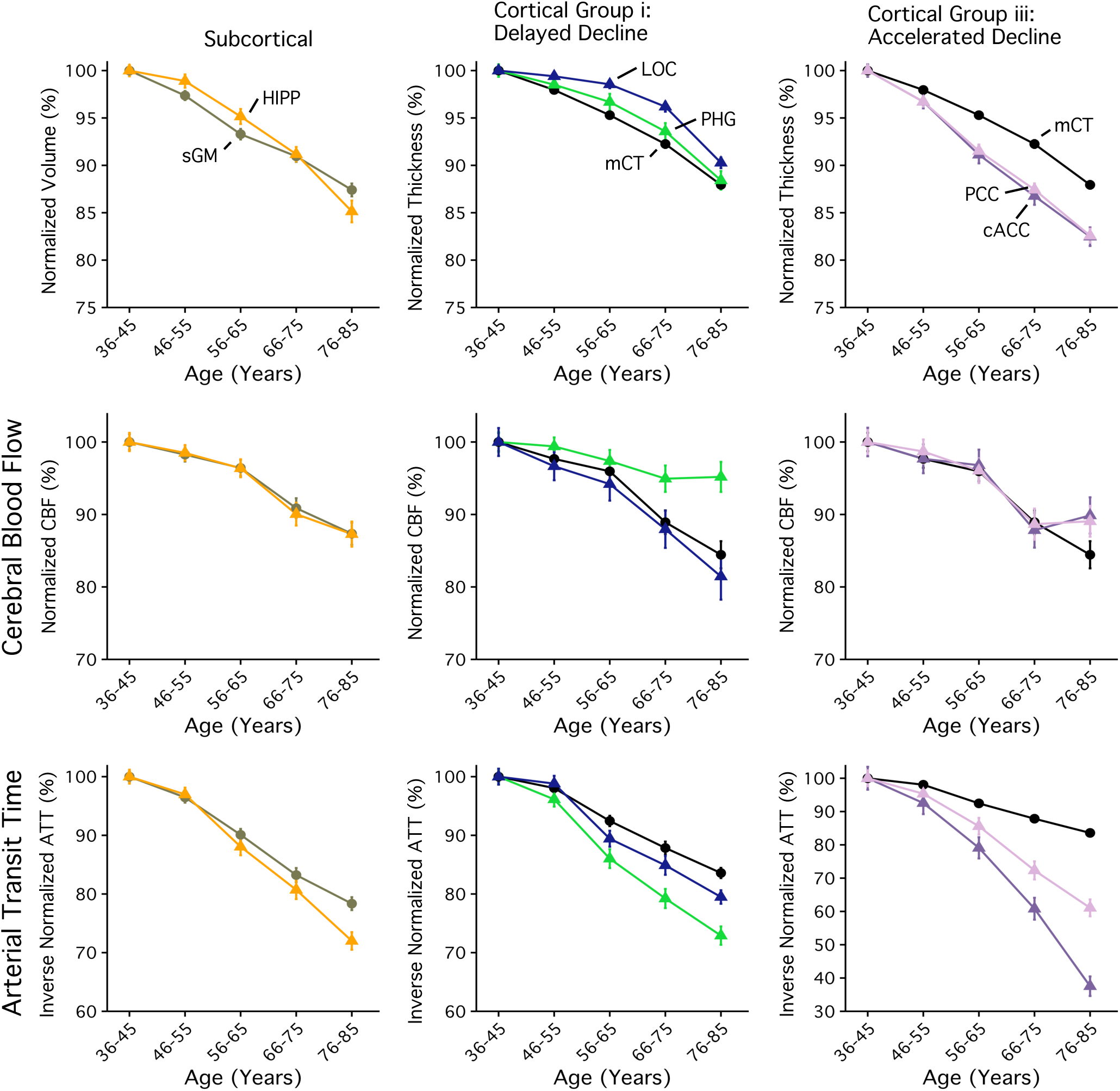
Age patterns from regional CBF and ATT are explored against patterns of accelerated and delayed decline seen from structural estimates in the HCP Aging cohort. The top row displays familiar patterns of *delayed* (i.e. HIPP volume and LOC and PHG thickness), and *accelerated* decline (i.e. cACC and PCC thickness) seen in the HCP Aging cohort and previously described in Figure 4. The middle row displays the normalized CBF, and the final row inverse normalized ATT. Regions showing accelerated decline also show nearly two times increase in ATT.

Age associations with CBF were comparable to those derived using structural estimates in regions representing *accelerated* and *delayed* decline groups. ATT increased nearly twice as fast in regions showing patterns of *accelerated* versus *delayed* decline.

Age associations obtained from structural MRI and ASL MRI for each region are displayed on the same axes in *Supplemental Materials 1.* Altogether, these results help to illustrate the relative strength of age associations from estimates obtained from T1-weighted images compared to estimates of CBF and ATT in the same region. Individual estimates of CBF or ATT compared to brain structure can also be found in *Supplemental Materials 1*.

### ATT shows the steepest associations with age

Finally, in Figure 6, the percent annualized rate (i.e. %/year) estimated from subcortical volume and regional cortical thickness were compared to those estimated from CBF and ATT for all the same regions. On average, annual rates of decline in subcortical regions obtained from volume-based estimates were comparable (range: −0.71-0.28%/year) to those from CBF (−0.29-0.02%/year) and ATT (−0.84-0.38%/year). Rates of decline across the cortex were on average comparable between regional thickness (−0.42-0.15%/year) and CBF (−0.73-0.14%/year), and twice as large when estimated from ATT (−1.64-0.27%/year)[F(1,25) = 11.3,p = 0.0025, b = 2.08, Rsq = 0.31]. In fact, regional ATT significantly predicted rates of decline in thickness (T = 3.37, p = 0.0025, Rsq = 0.28), even when including CBF as a covariate (T = 3.48, p = 0.002, Rsq = 0.29).

**Figure 6.**
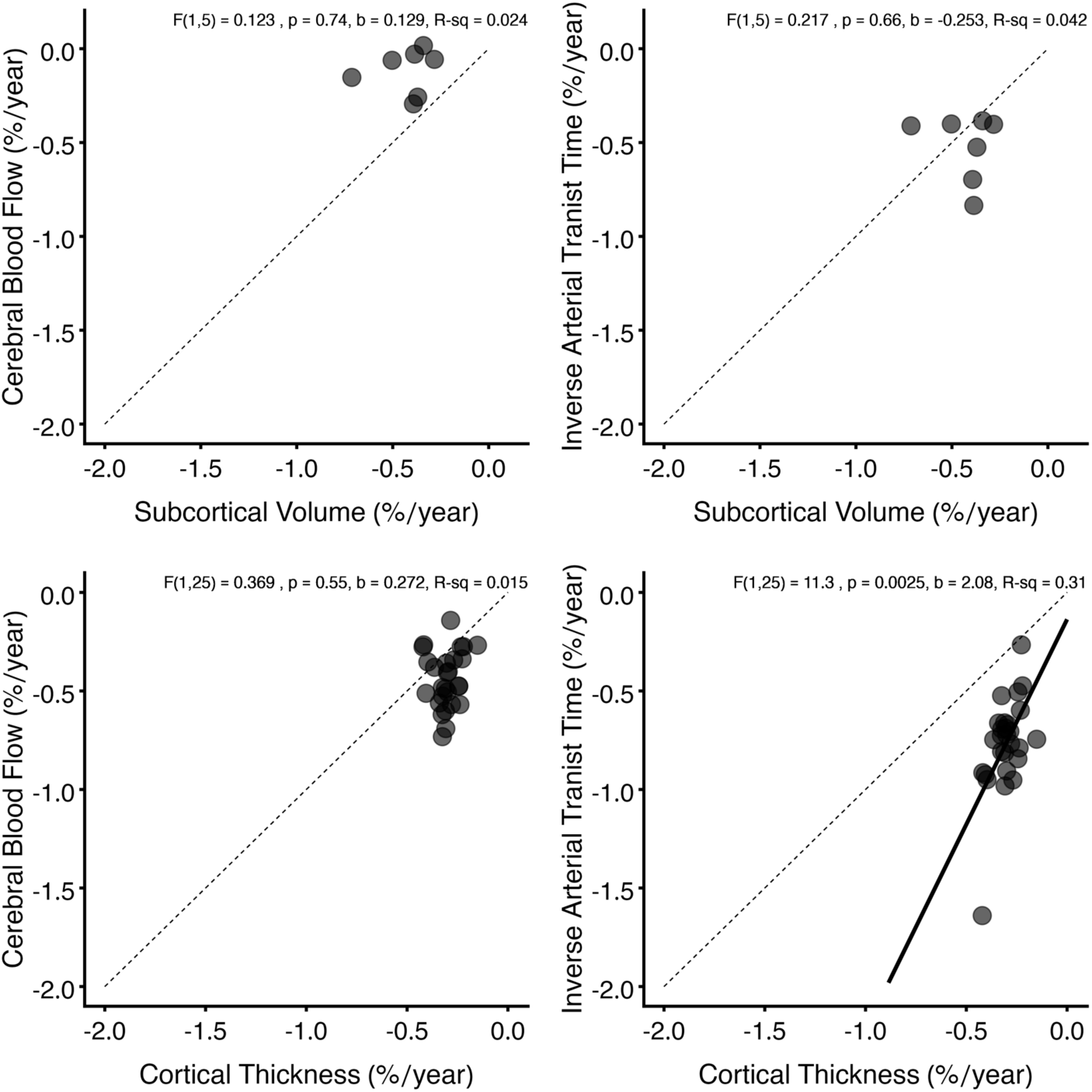
Across the cortex, regional age associations were the greatest when estimated by ATT from ASL images and map to delayed, steady and accelerated decline groups. Age associations, represented here as the approximate percent annualized rate (%/year), estimated from subcortical volumes (top row) and regional cortical thickness (bottom row) are plotted against estimates of CBF (left side), and ATT (right side) from the same regions. Regions falling along the identity line would show comparable age associations between measures. Subcortical volume-based estimates provided greater age associations (range: −0.71-0.28%/year) as compared to those estimated from CBF in the same regions (range: −0.29-0.02%/year). Cortical regions showed the greater age associations with inversed normalized ATT (range: −1.64-0.27%/year) compared to those from thickness estimates (range: −0.42-0.15%/year).

## Discussion

Our findings reveal recurring patterns of *accelerated* and *delayed* decline across the adult lifespan that were (1) present across subcortical and cortical structures, (2) replicated in two large, independent datasets, and finally (3) across two imaging modalities. Notably, these spatial trajectories mirrored the organization of major arterial flow territories, suggesting that vascular architecture may serve as a key organizing principle in brain aging.

### Regional Patterns of Accelerated, Steady and Delayed Decline

Regions such as the medial prefrontal and cingulate cortices showed *accelerated* decline, consistent with their heightened metabolic and vascular demand ^12,20,49–52^. In contrast, medial temporal regions including the hippocampus and entorhinal cortex exhibited *delayed* decline, in line with previous reports ^6–8,21–23^. These regions subserve memory and cognitive flexibility and represent early sites of neurodegenerative vulnerability ^53–63^. Apart from the PCC, regional heterogeneity emerged consistent across datasets (HCP Aging and UK Biobank), which underscores the stability and potential biological relevance of these vascular aging patterns. One possible explanation for why decline in the PCC was not replicated could in part be due to the slightly narrowed window across the adult lifespan, and may further imply that the peak or maximal rate of decline in the PCC is earlier in adulthood. Interestingly, separation between cACC and PCC was also found with estimates of ATT in the HCP Aging cohort, where rates of ATT increased for cACC faster than for PCC (see Figure 5). These differences should be further explored.

### Vascular Topography as an Organizing Principle

A major advance of this work is the demonstration that regional aging trajectories align with the brain’s vascular territories. Our data-driven clustering approach, independent of any vascular priors, recapitulated the boundaries of the anterior, middle, and posterior cerebral arteries. This alignment suggests that the anatomy of vascular supply exerts a strong influence on how aging unfolds spatially. Perfusion heterogeneity, local metabolic demand, and chronic vascular stress may interact to produce distinct rates of decline across arterial territories ^24,26,28^. This interpretation is consistent with evidence of vascular remodeling in aging, including vessel rarefaction, increased tortuosity, reduced elasticity, and endothelial senescence ^30,31^. Such microvascular deterioration precedes measurable gray matter loss, hinting that vascular aging is an upstream driver of morphological change. Together, these multiscale processes establish vascular architecture as both a constraint and a catalyst of age-related brain decline.

### Value of Multimodal Imaging

The combination of ASL-derived ATT and structural MRI provided convergent evidence that vascular transit slowing parallels tissue loss. While CBF showed comparable associations with age to structural estimates, inverse ATT exhibited robust age associations changes that mirrored those seen in structural decline. In fact, inverse ATT showed steeper age associations in nearly all estimates examined. Unlike structural MRI, which captures tissue loss, ATT reflects vascular transit dynamics that may change prior to overt atrophy. These findings echo prior work showing that perfusion inefficiency and blood–brain barrier dysfunction emerge early during healthy brain aging and likely contribute to later cognitive decline ^49,51,64–66^ At the microstructural level, losses in capillary density and neurovascular coupling could degrade perfusion efficiency and hamper metabolic support, while at the macro level, arterial stiffness and flow variability could amplify vulnerability within specific territories. Multimodal imaging therefore offers a bridge between physiology and structure, supporting hypotheses of vascular primacy in the aging process.

### Study Limitations

First, we acknowledge the inherent limitations in making inferences about aging from cross-sectional populations. Cross-sectional designs, while useful, likely under-estimate individual rates of change ^10,18^, and do not fully capture the dynamic changes that occur within an individual over time ^67^. However, such cohorts can and have already provided valuable insight for classification of whole-brain age based on biological age ^68,69^, neurodegenerative disorders ^57,68^, psychiatric disorders ^70,71^, or to evaluate/identify key lifestyle factors contributing to the aging process ^72^. Second, an unexpected association between total intracranial volume and age was observed in our sample and was mitigated after adjustment using the atlas transform error term (see *Appendix 1*). However, we acknowledge this adjustment does not fully resolve all issues with registration of individual estimates to standard space. Third, the use of atlas-based measures may have limited our ability to resolve some regional boundary disputes for arterial supply. As noted previously, three regions defined using the Desikan-Killiany atlas – the precuneus, superior frontal and superior parietal – fell nearly equally along flow territories and were excluded. Some subcortical structures may also benefit from non-atlas-based approaches. For example, the ventral nuclei of thalamus are supplied by MCA, while dorsal portions are supplied by the PCA ^45^. Moreover, the ventral striatum was incorrectly defined as being predominantly supplied by the MCA, however, all previous work suggest it is supplied by the ACA ^41,42^. The corrected assignment was used in this study. Defining individual-based maps of arterial flow territories would provide the greatest accuracy and remove spatial heterogeneity due to individual differences from vascular architecture ^43^.

### Implications and Future Directions

Our findings extend the growing view that vascular health is central to both normative and pathological brain aging. The identification of territory-specific decline highlights that vascular topology imparts spatial constraints on the aging process. Beyond its descriptive value, this spatial framework presents a mechanistic avenue for linking vascular risk, perfusion, and atrophy patterns. Importantly, regional patterns could serve as a promising biomarker in longitudinal studies and clinical trials targeting age-related or vascular-related brain decline.

Future work should integrate individualized vascular mapping, longitudinal follow-up, and direct assessment of endothelial and metabolic function to clarify whether changes in vascular efficiency or metabolic demand drive the observed patterns. Such avenues could reveal novel vascular biomarkers of early aging and aid in designing interventions that target perfusion maintenance or vascular elasticity.

## Conclusions

To our knowledge, this is the first study to demonstrate that regional-specific age trajectories across the brain align with the arterial flow architecture that underpins it. These findings highlight that vasculature is not simply a supportive framework but a dynamic and organizing determinant of structural and physiological aging. Continued integration of multimodal imaging and physiological metrics will be essential for clarifying the causal role of vascular health in preserving brain function across the lifespan.

## Data Availability

All data are available online

## Acknowledgements

Research reported in this publication was supported by the National Institute On Aging of the National Institutes of Health under Award Number U01AG052564 and by funds provided by the McDonnell Center for Systems Neuroscience at Washington University in St. Louis. The HCP-Aging 2.0 Release data used in this report came from DOI: <u>10.15154/1520707</u>. Additionally, this research was supported by the NIH-NIDCD grant K23DC019179, and A2 Collective grant by MassArt (PI: Marziye Eshghi). The UK Biobank analyses were conducted under application number #32568. We thank John Jacobi for his role in optimization of HCP ASL data processing.

## Author contributions

L.C.H. had a major role in study conceptualization, data analyses, interpretation of the findings, visualization, and writing of the manuscript. M.E. contributed to the study conceptualization, interpretation of the findings, and manuscript preparation and provided feedback. J.D. contributed to data analyses, interpretation of the findings, visualization and provided feedback. R.L.B contributed to the initial study conceptualization, funding, data analyses, interpretation of findings, visualizations, and provided feedback. R.W.M. contributed to data analyses, interpretation of the findings, and provided feedback. T.G. provided feedback. M.R.J. had a major role in the study conceptualization, data analyses, interpretation of the findings, visualizations, manuscript preparation and provided feedback. D.H.S. had a major role in the study conceptualization, data collection, funding, data analyses, interpretation of the findings, visualizations, manuscript preparation and provided feedback. All authors contributed to the article and approved the submitted version.

## Appendix 1 – reprinted from Hanford et al., 2025 Human Brain Mapping

### Adjusted estimated Total Intracranial Volume (eTIV)

Volume-based structural measures are confounded by head size, and are typically corrected for using estimated Total Intracranial Volume (eTIV). While eTIV should remain consistent across the lifespan (Buckner et al., 2004), it was noted to have a negative association with age in the HCP-Aging dataset. This age-association was diminished (for female participants) and removed (for male participants) after adjusting eTIV by FreeSurfer’s atlas transform error score (atlas_txfm_error) using the following equation:

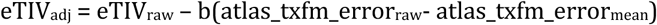

where eTIV_adj_ is the adjusted measure, eTIV_raw_ is the automated measure provided by FreeSurfer’s recon-all pipeline, b is the slope of the atlas transform error with age, atlas_txfm_error_raw_ is the individual measure and atlas_txfm_error_mean_ is the average atlas transform error across the HCP-Aging sample.

All reference to eTIV in the manuscript is to this adjusted measure of eTIV. Adjusted eTIV was used to correct for head size in volume-based estimates (including hippocampal volume and total gray matter volume).

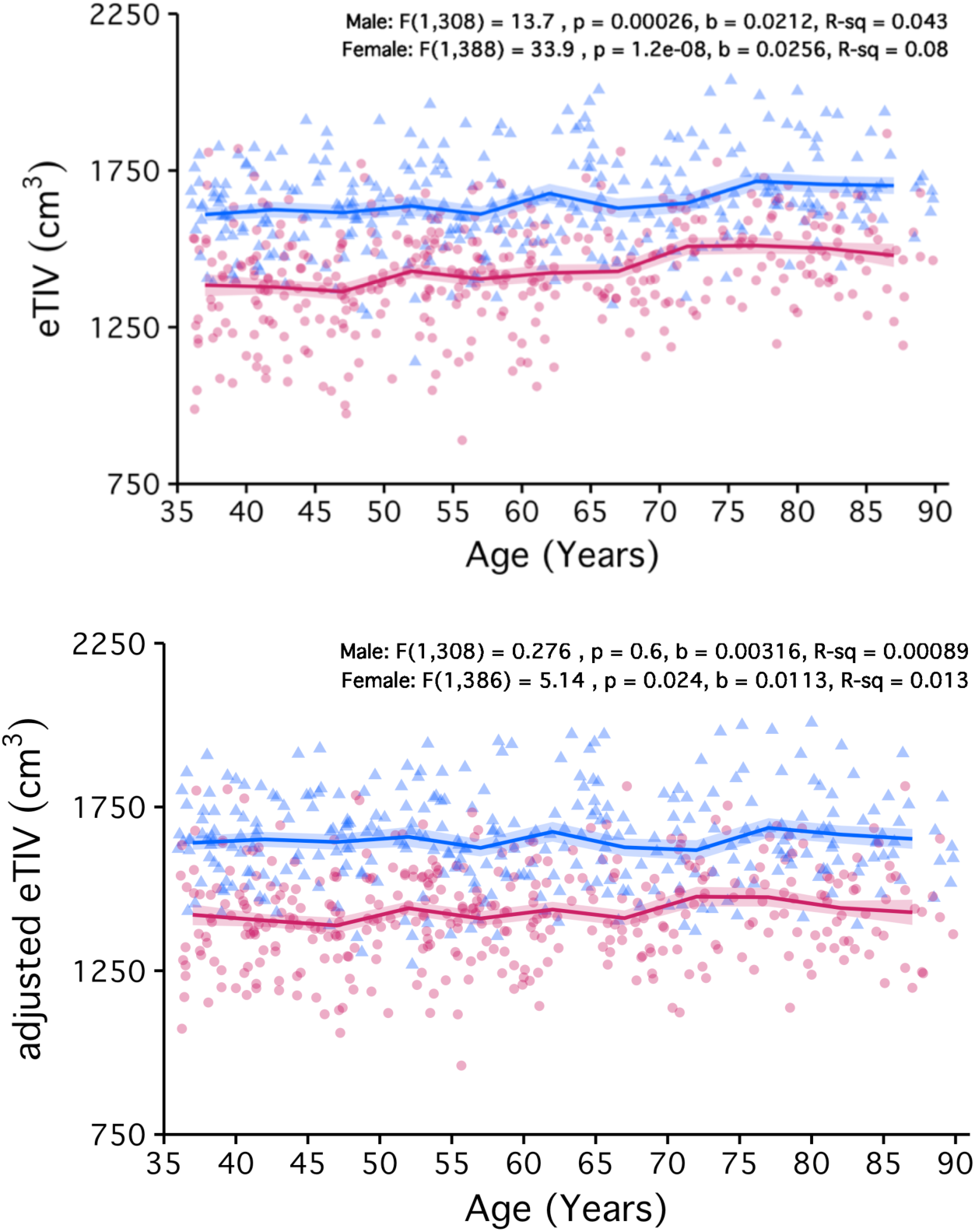

**Figure S1.**
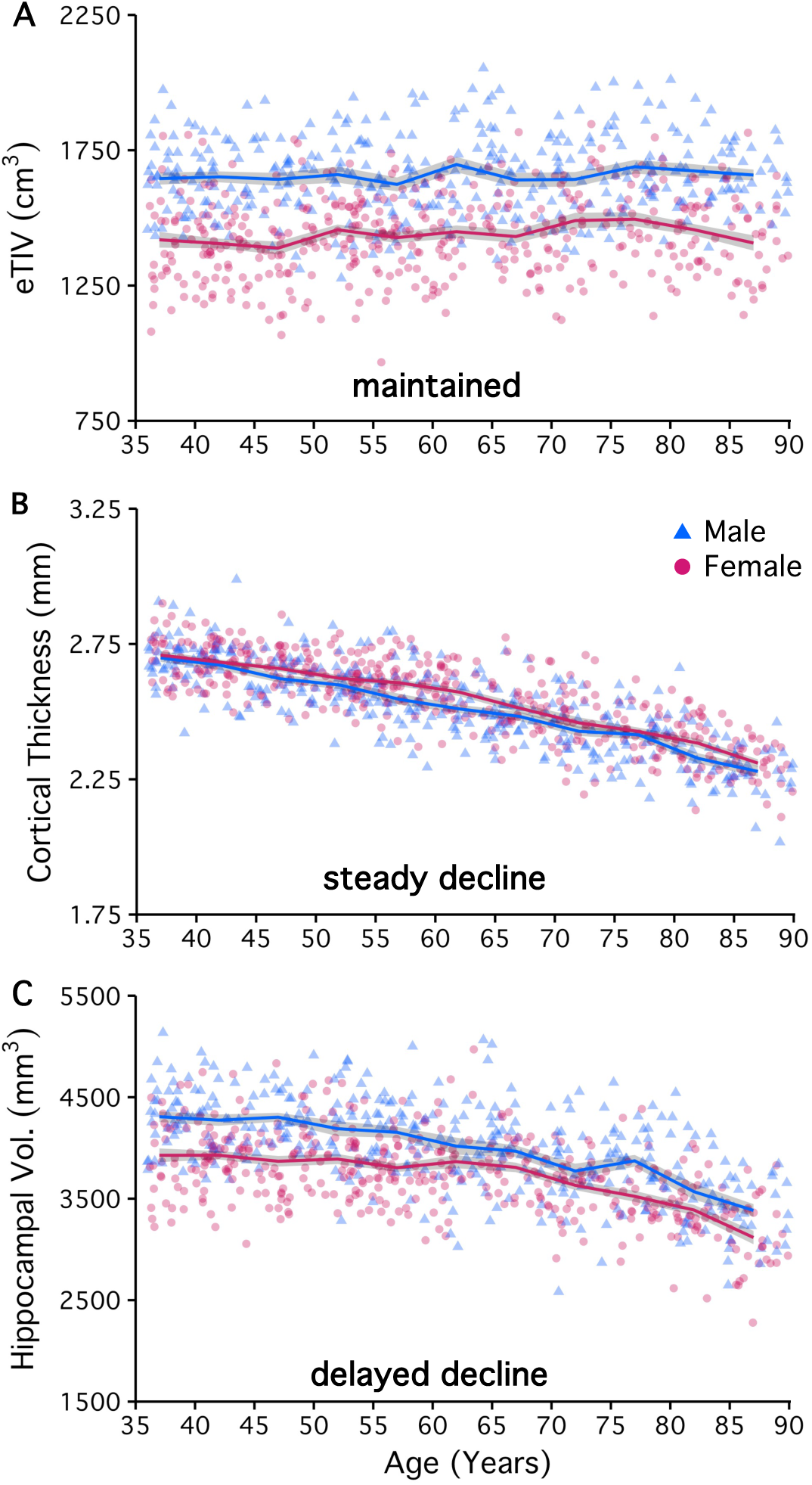

**Figure S2.**
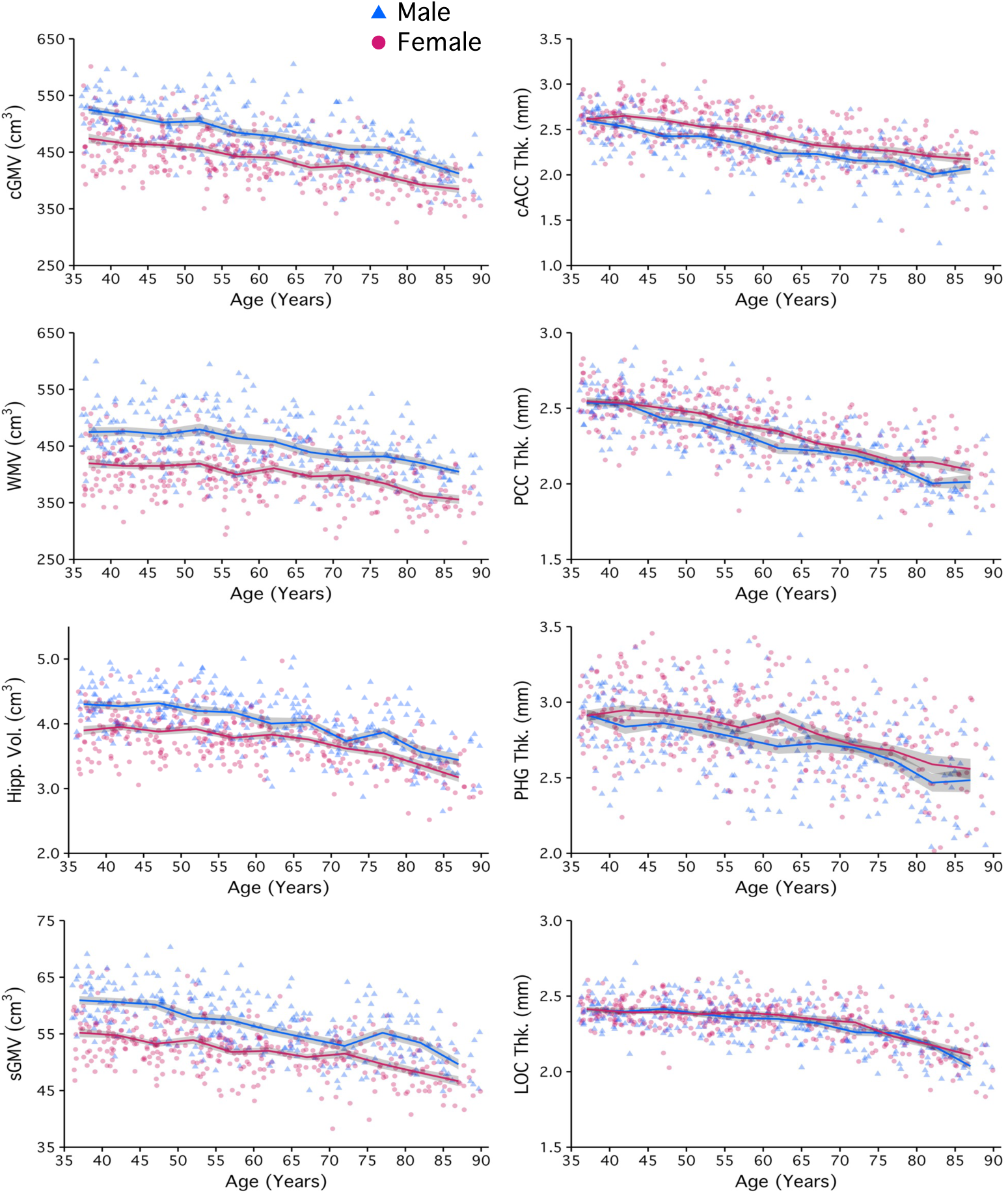

**Figure S3.**
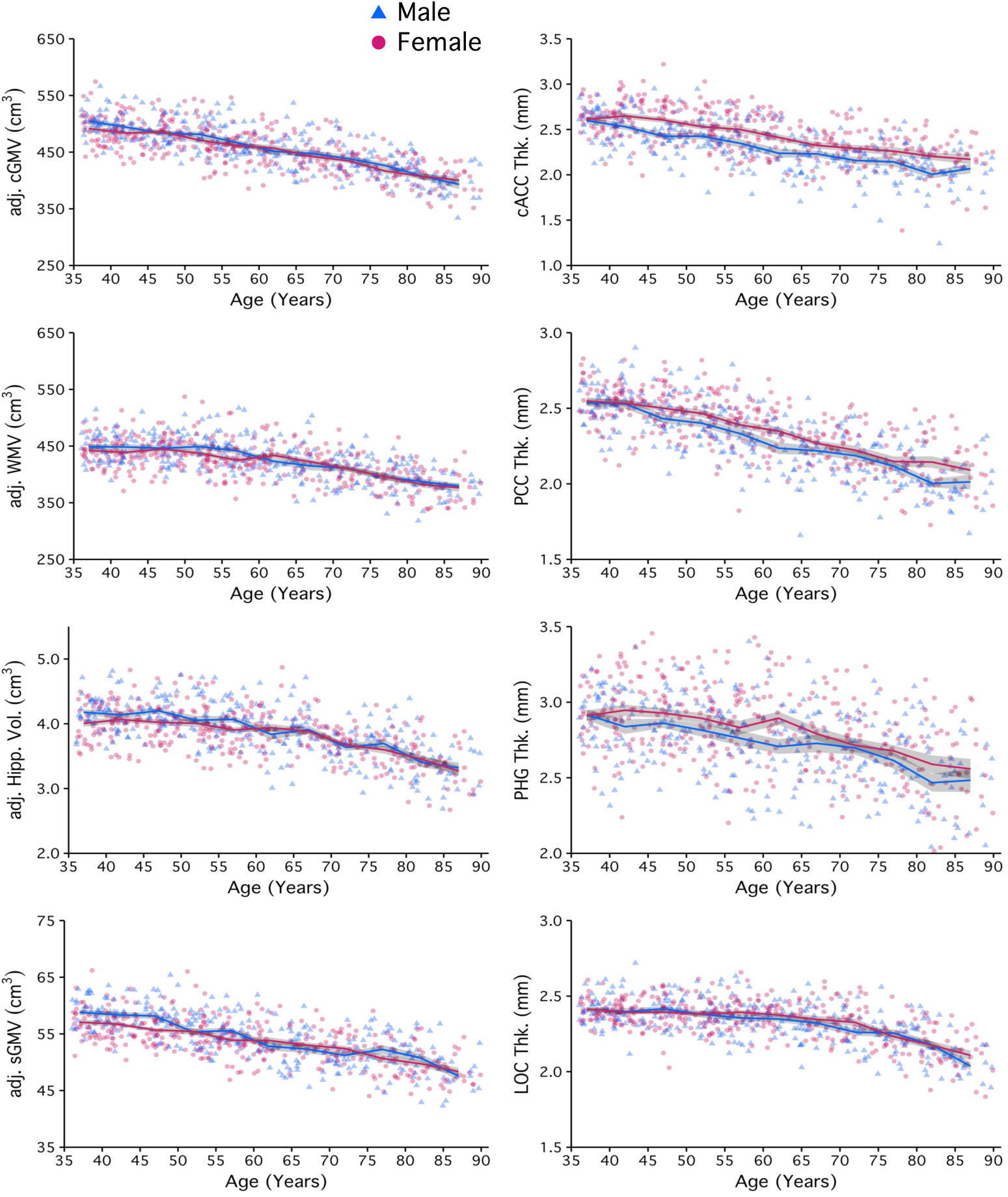

**Figure S4.**
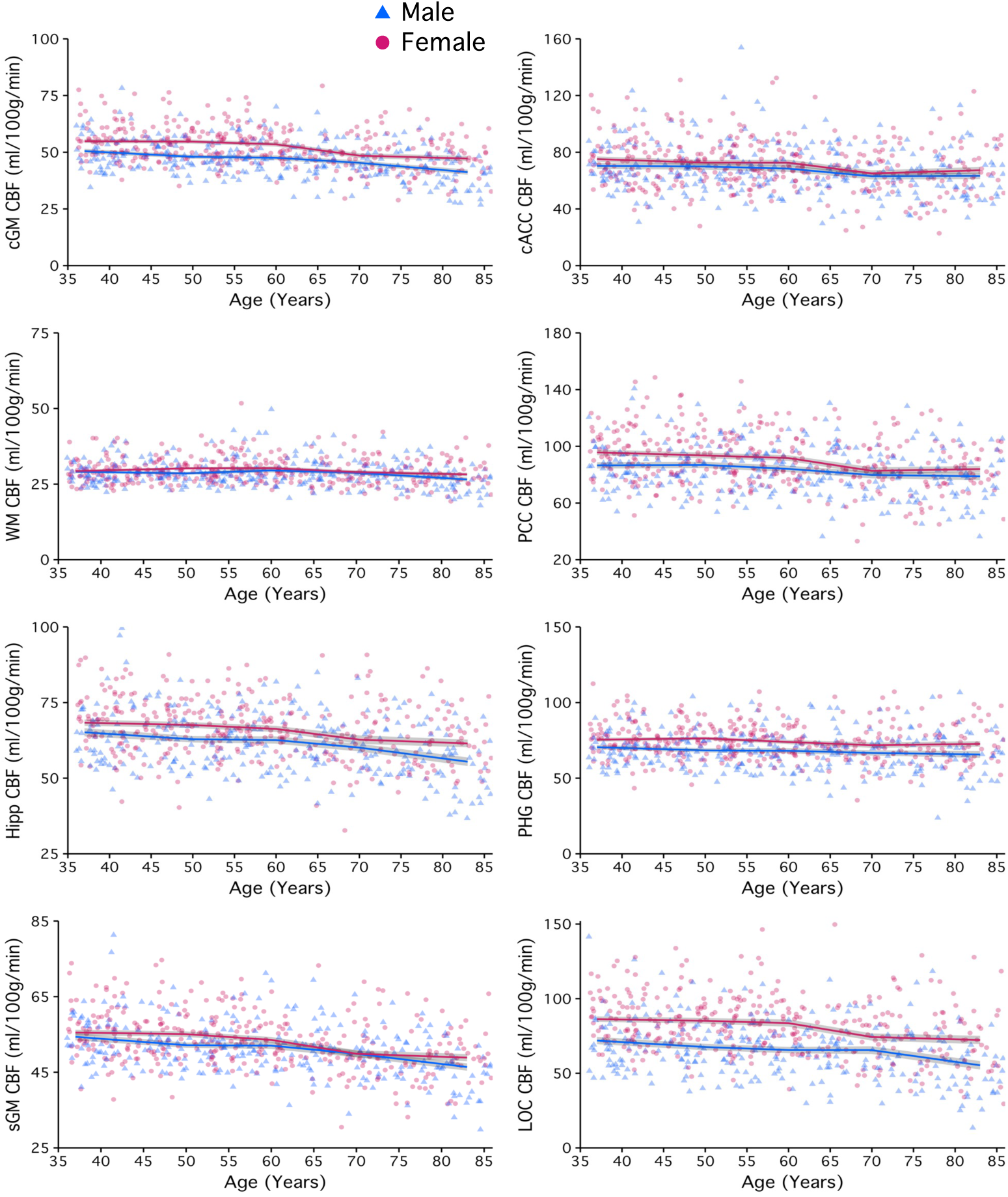

**Figure S5.**
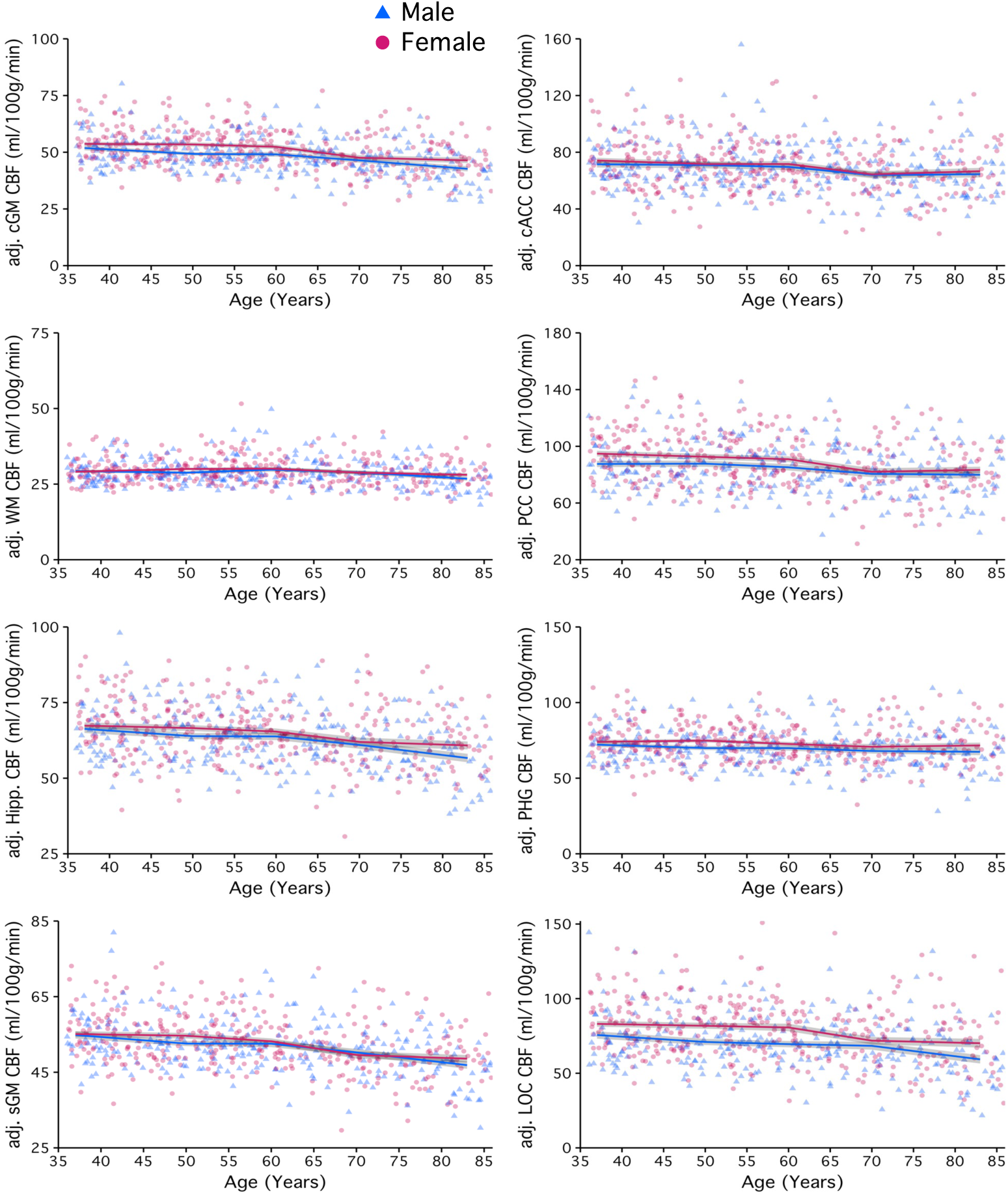

**Figure S6.**
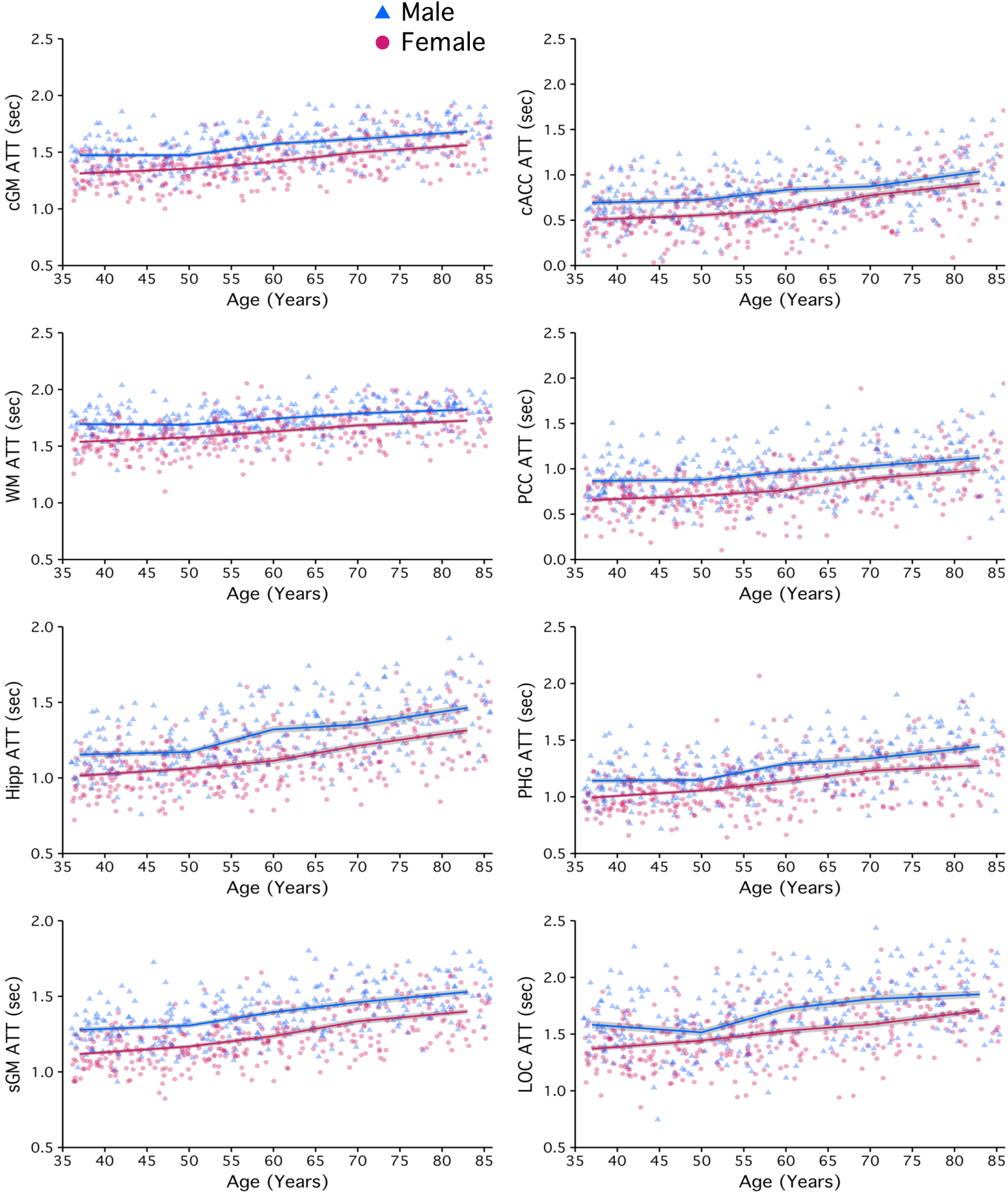

**Figure S7.**
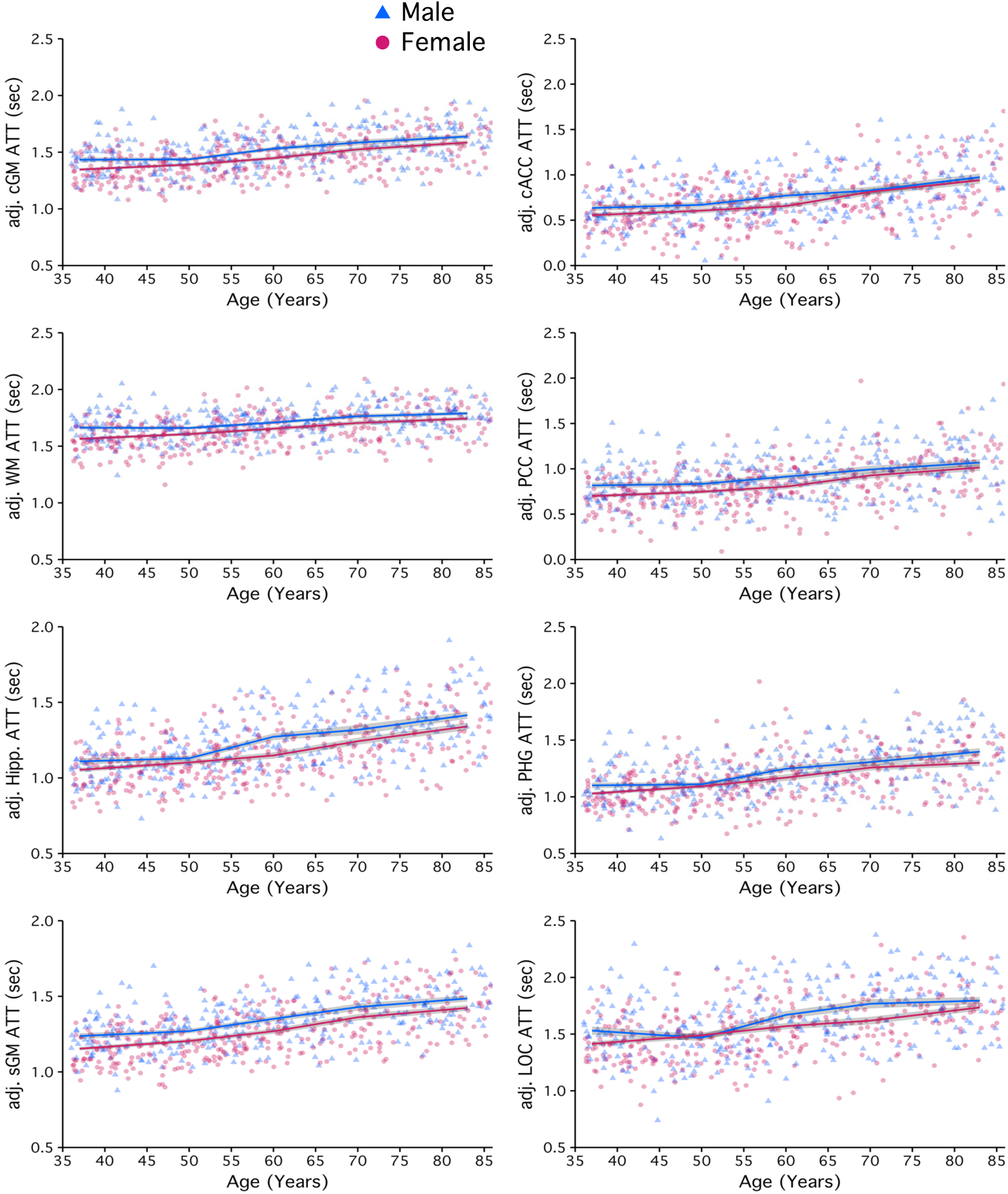

**Figure S8.**
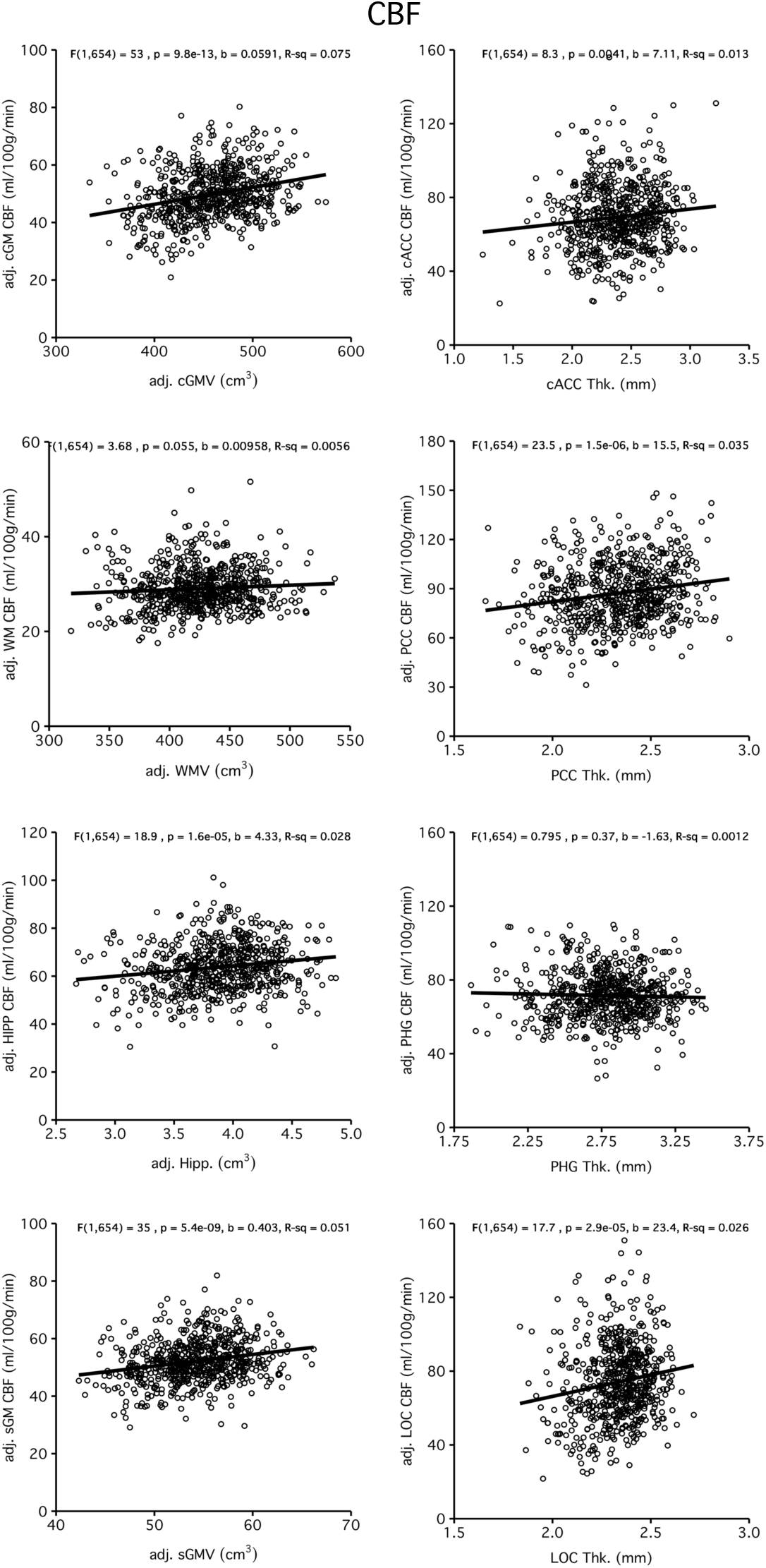

**Figure S9.**
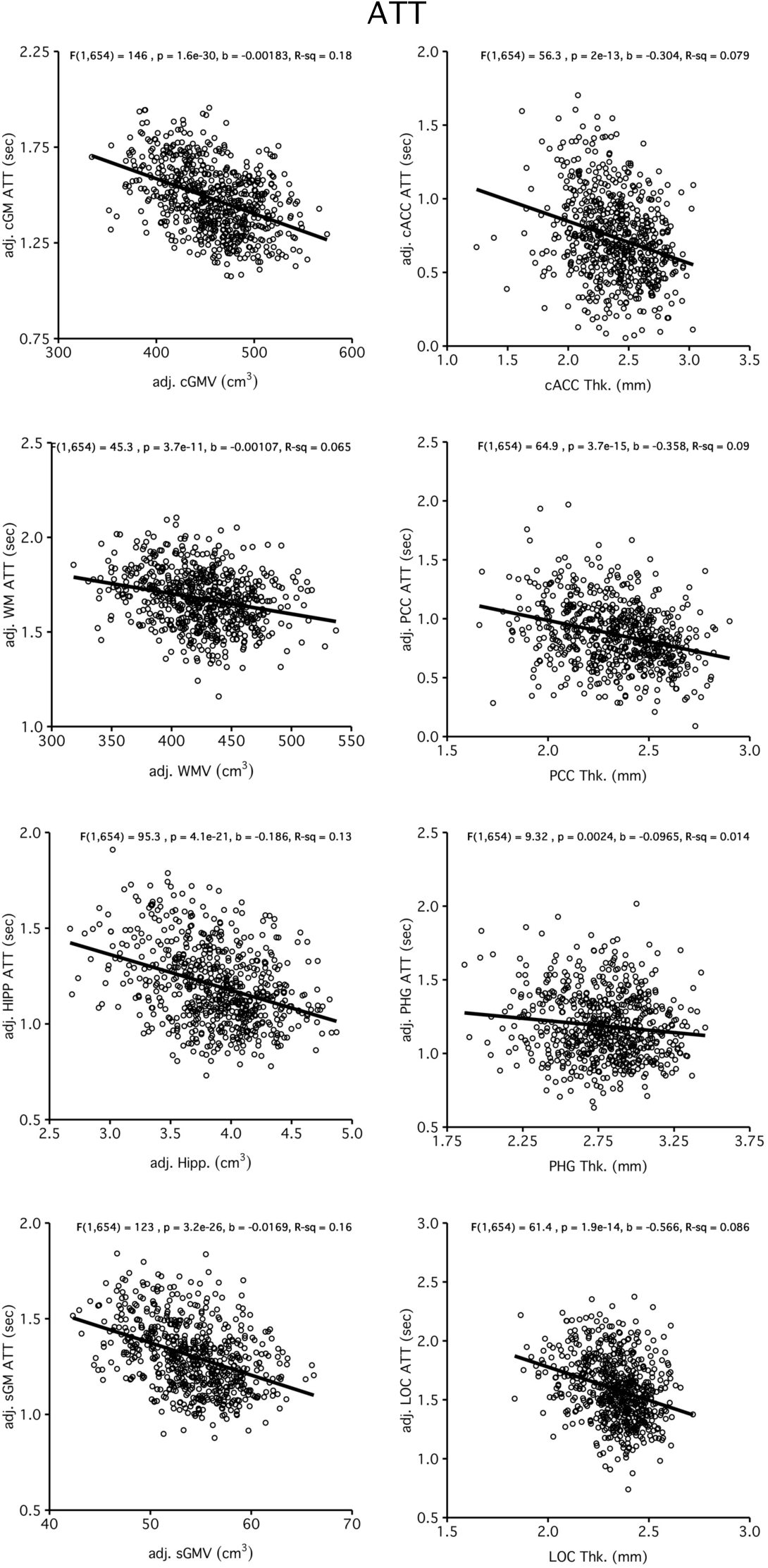

**Figure S10.**
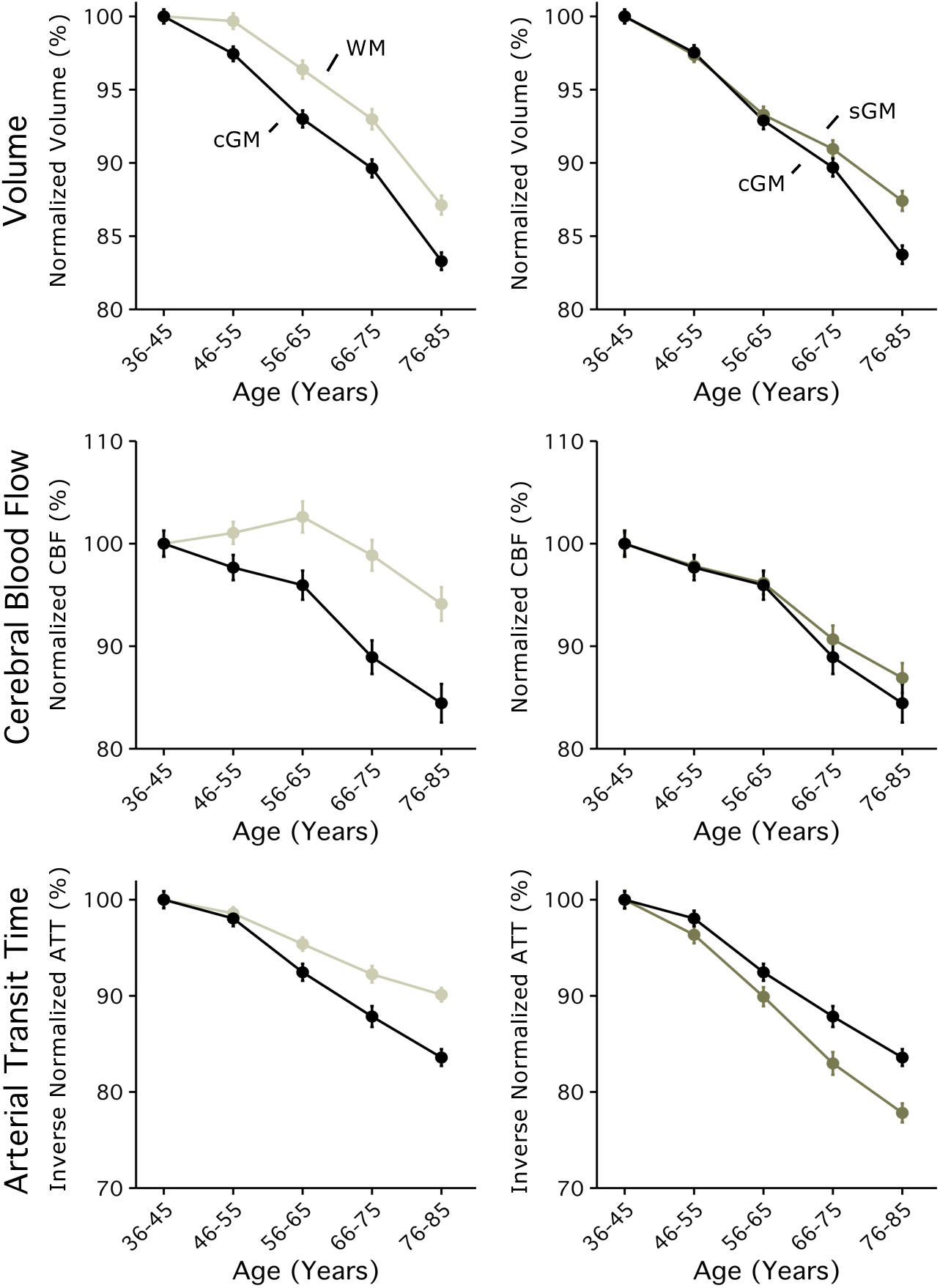

**Figure S11.**
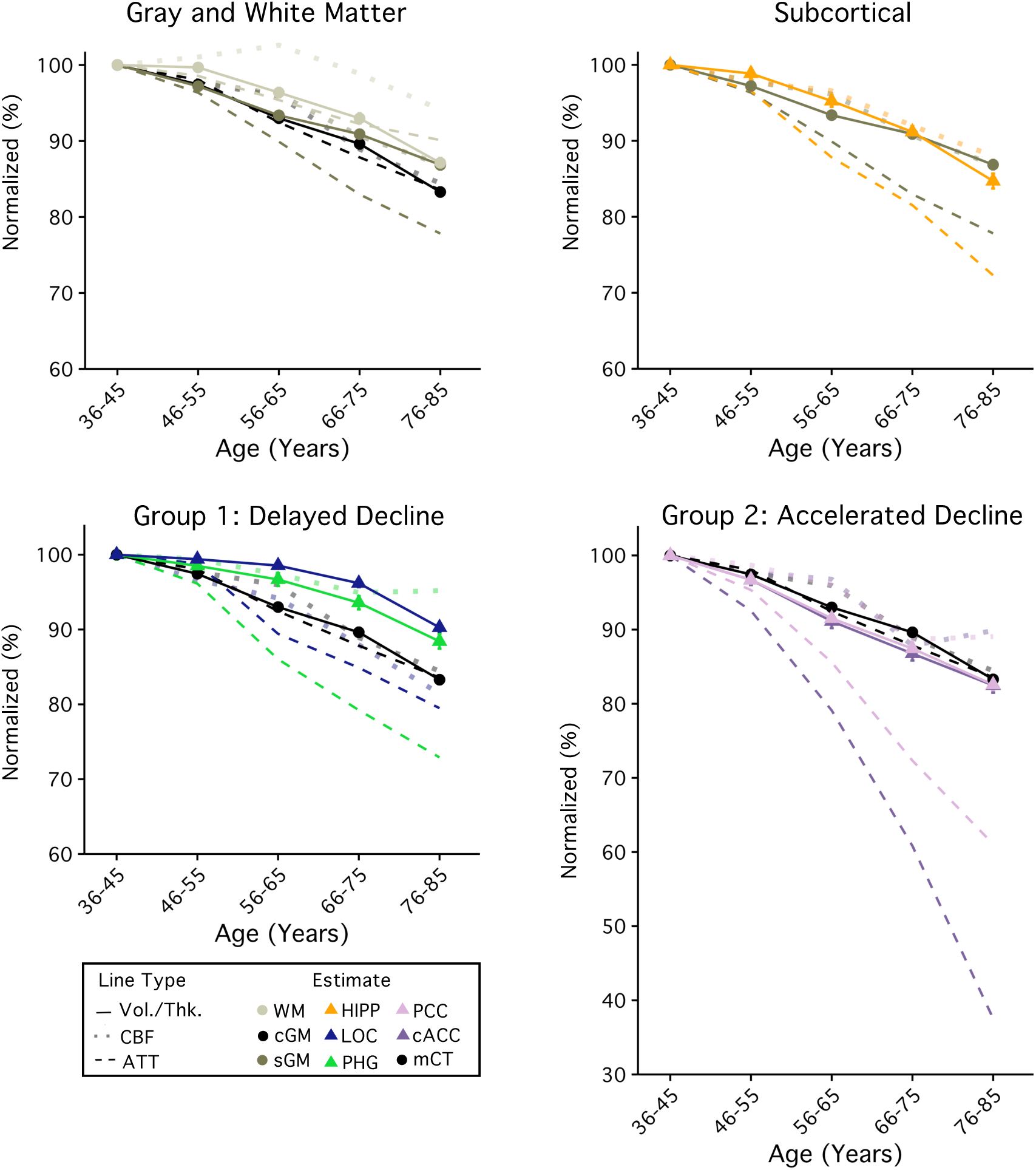

## Supplemental Material 2: Reliability Sample Methods and Results

### Methods

#### Overview

The aim of this sub-study was to better characterize the reliability of regional estimates at a group size that reflected the smallest age group in our analysis (n=31). In doing so, we sought to better understand the limitations of sample size in our analysis. Participants completed two scan sessions within the same week; where an HCP-protocol T1-weighted (T1w) image was acquire at each timepoint. Test-retest reliability was assessed by comparing regional measures estimated from each of the T1w images acquired on separate days (Day 1 vs Day 2).

#### Participants

Participants were 31 healthy adults, aged 19-42 years [(mean = 26.1 ± 5.3 years), 17 were female] recruited from the local Harvard community (e.g., student, staff, or fellow). Participants were free from COVID symptoms, any neurological, psychological, or other MR contraindications. Written informed consent was given and participants were paid for the time.

#### Image Acquisition, Processing and Analysis

Structural MR scans were acquired at the Harvard Center for Brain Science using a 3T Siemens Magnetom Prisma-fit MRI scanner (Siemens Healthcare, Erlangen, Germany), and 32-channel head coil. T1w images were an 0.8 mm multi-echo MPRAGE (van der Kouwe *et al.* 2008); the same HCP-imaging protocol described in the manuscript. Images were processed using standard automated pipeline from FreeSurfer v6.0.0 (Dale *et al.* 1999; Fischl *et al.* 1999). Reliability was calculated for all estimates (i.e. 17 subcortical volumes and 68 regional cortical thickness) as the amount of shared variance across days (R^2^ coefficients from linear regression models comparing Day 1 vs Day 2). Reliability from subcortical volumes were displayed in relation to their size (mm^3^), and regional thickness estimates were relative to their surface area (mm^2^). Estimates were excluded if either left/right hemisphere estimates showed poor reliability (R^2^ <0.6).

### Results

#### Most subcortical volume and regional cortical thickness estimates are highly stable

Subcortical regions showing the greatest permanence across timepoints included hippocampal volume (L/R HIPP R^2^ = 0.92/0.92)(see Supplemental Figure 1a), brainstem (BST R^2^ = 0.98), putamen (PUT: R^2^ = 0.94/0.89), and ventral diencelphon (vDC: R^2^ = 0.86/0.88). Even smaller regions including the ventral striatum (VS: R^2^= 0.83/0.71), and amygdala (AMYG: R^2^ = 0.74/0.76) and pallidum (PALL: R^2^= 0.78/0.73) met our minimum criteria and were included. Reliability of all subcortical volumes are plotted by size in Figure 1B.

Estimates of regional cortical thickness were observed to have higher reliability included parahippocampal volume (PHG R^2^ = 0.94/0.88)(See Figure 1C), caudal middle frontal (cMFG R^2^ = 0.91/0.85), inferior parietal (IPC R^2^ = 0.84/0.83), fusiform (FG R^2^ = 0.87/0.81), lateral occipital (LOC R^2^ = 0.88/0.85), paracentral (ParaC R^2^ = 0.96/0.90), inferior frontal (IFGoper R^2^ = 0.92/0.83; IFGorb R^2^ = 0.95/0.85; IFGtri R^2^ = 0.87/0.84), Pericalcarine (CAL R^2^ = 0.86/0.86), precunenus (PreCUn R^2^ = 0.90/0.84), rostral middle frontal (rMFG R^2^ = 0.89/0.85), and superior frontal (SFG R^2^ = 0.91/0.90), supramarginal (SMG R^2^ = 0.84/0.91), and superior temporal (STG R^2^ = 0.92/0.87). Regions showing poor reliability included the medial orbitofrontal (mOFC R^2^ = 0.33/0.85), lateral orbitofrontal (lOFC R^2^ = 0.64/0.38), insular (INS R^2^ =0.16/0.37), precentral (PreC R^2^ = 0.59/0.69), Bank of the superior temporal sulcus (bankSTS R^2^ =0.54/0.83), rostral anterior cingulate (rACC R^2^=0.58/0.80) as well as temporal pole (TP R^2^ =0.64/0.55). These regions (n=6) were excluded from further analysis.

**Figure S2-1.**
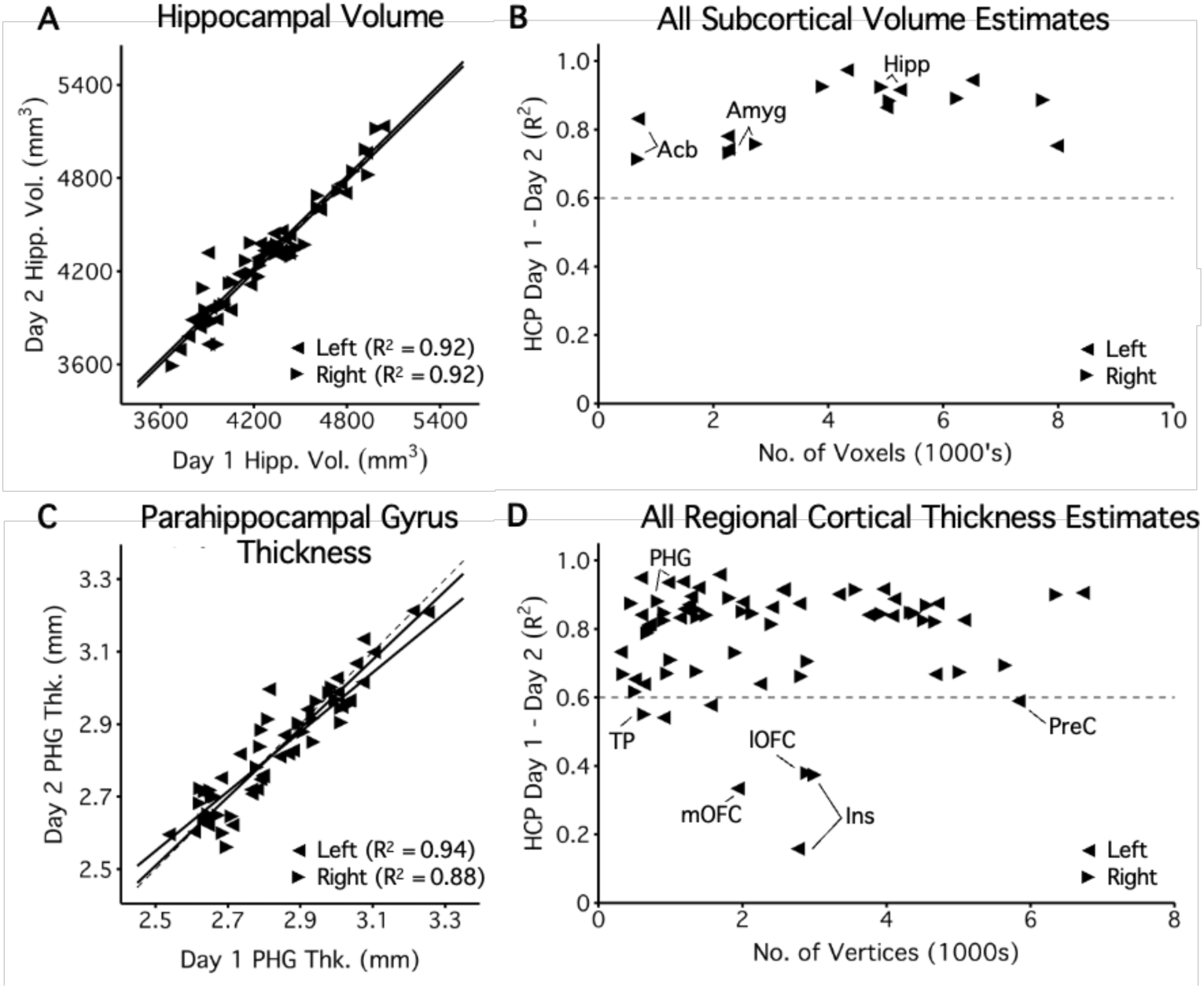
Reliability of regional subcortical volume and cortical thickness estimates from an independent sample of 31 healthy adults.

Finally, while caudate volume appeared reliable from this sub-study (CAUD R2 = 0.97/0.92), it was excluded from our analyses as it displayed a “biologically implausible” association with age that defied any reasonable interpretation (see Figure 2).

**Figure S2-2.**
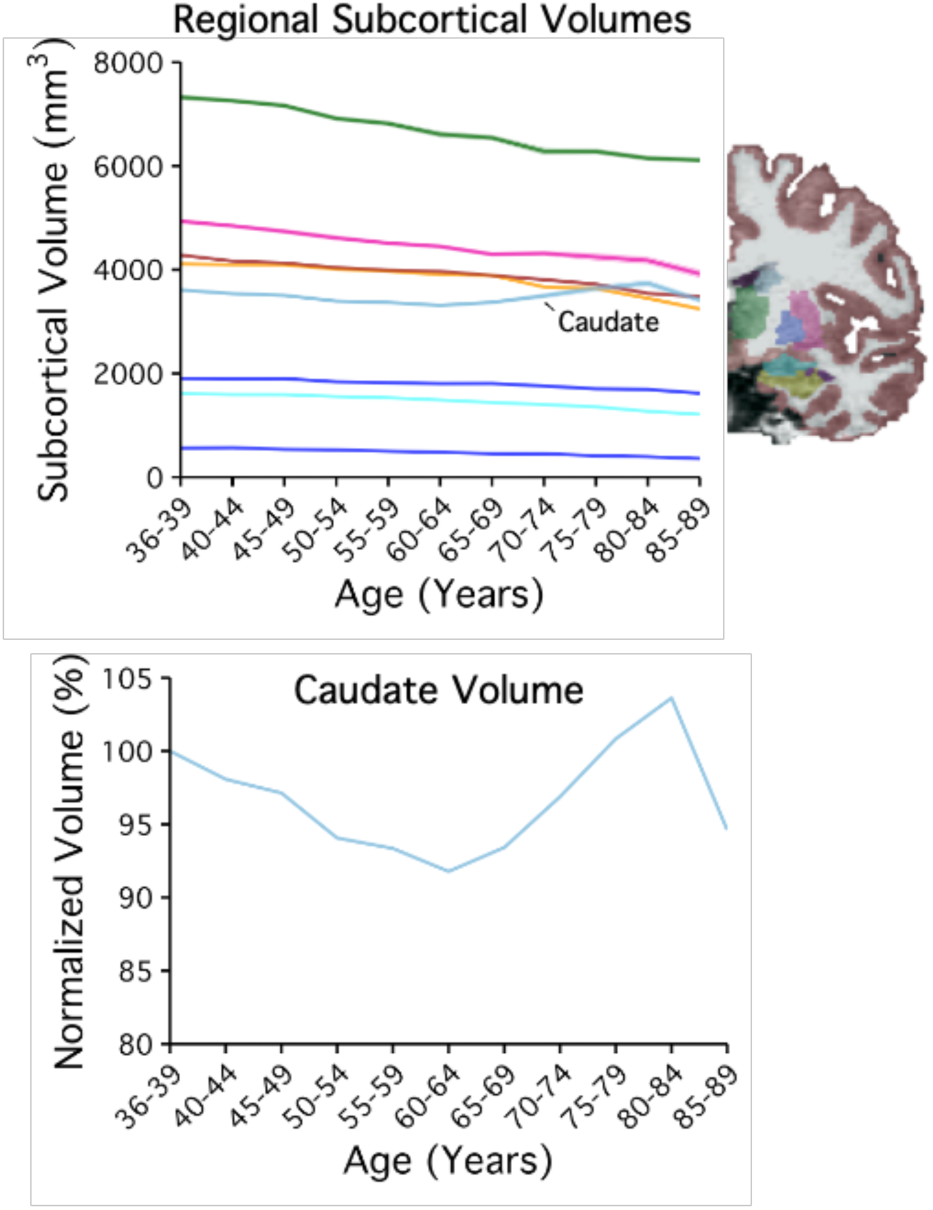
Subcortical volumes across the adult lifespan. Normalized caudate volume with age.

